# What is the long-term impact of COVID-19 on the Health-Related Quality of Life of individuals with mild symptoms (or non-hospitalised): A rapid review

**DOI:** 10.1101/2022.09.09.22279642

**Authors:** Llinos Haf Spencer, Annie Hendry, Abraham Makanjuola, Jacob Davies, Kalpa Pisavadia, Dyfrig Hughes, Deb Fitzsimmons, Clare Wilkinson, Rhiannon Tudor Edwards, Ruth Lewis, Alison Cooper, Adrian Edwards

**Affiliations:** Bangor Institute for Health and Medical Research, Bangor University; Swansea University; Wales COVID-19 Evidence Centre

## Abstract

The COVID-19 morbidities model has been widely used since 2020 to support Test and Trace and assess the cost-effectiveness of the COVID-19 vaccination programme. The current iteration of the Long COVID model covers several morbidities associated with COVID-19, which are essential to plan for elective care in the future and identify which services to prioritise. However, there are uncertainties in the model around the long-term health-related quality of life (HRQoL) impact of COVID-19, which is primarily based on data for severe COVID disease or hospitalised patients at present. The COVID-19 morbidities model requires updating to address gaps and reflect the latest HRQoL evidence.

The aim of this rapid review was to provide updated HRQoL evidence for the COVID-19 morbidities model to better support decision-making in relation to COVID-19 policy.

Thirteen primary studies were identified. People who had an initial mild COVID-19 illness or were not treated in hospital can have a decreased HRQoL post-COVID. However, the extent, severity, and duration of this is not consistent. The evidence on the long-term impact of a mild COVID-19 infection on HRQoL is uncertain.

Implications for policy and practice include:

An initial mild COVID-19 illness can lead to a reduction in HRQoL and impaired mental health, but there is evidence indicating that patients can show significant recovery up to normal levels after one year.
Employers should be aware that employees may have prolonged experiences of impaired mental health, including anxiety, depression, and fatigue, following COVID-19 disease, even if their initial disease was mild (not hospitalised).
Public health agencies should make patients with mild COVID-19 disease aware of the potential for ongoing symptoms and ways to mitigate and manage them through raised awareness and education.
Health Boards should review their provision of long-COVID services in relation to the extent of impacts identified.
Better quality studies that report longitudinal follow-up data on HRQoL for a representative cohort of patients who have had mild COVID-19 are required.

**Funding statement:** The Bangor Institute for Health and Medical Research, Bangor University was funded for this work by the Wales Covid-19 Evidence Centre, itself funded by Health & Care Research Wales on behalf of Welsh Government.

**Rapid Review Details:** *Review conducted by:* Bangor Institute for Health and Medical Research Rapid Review Team

*Review Team:*

- Dr Llinos Haf Spencer,
- Dr Annie Hendry,
- Mr Abraham Makanjuola,
- Mr Jacob Davies,
- Miss Kalpa Pisavadia,
- Professor Dyfrig Hughes,
- Professor Deb Fitzsimmons,
- Professor Clare Wilkinson,
- Professor Rhiannon Tudor Edwards

*Review submitted to the WCEC in:* July 2022

*Rapid Review report issued by the WCEC in:* August 2022

*WCEC Team:* Adrian Edwards, Ruth Lewis, Alison Cooper and Micaela Gal involved in drafting, Topline Summary, editing etc.

*This review should be cited as:* RR00040. Wales COVID-19 Evidence Centre. What is the long-term impact of COVID-19 on the Health-Related Quality of Life of individuals with mild symptoms (or non-hospitalised): A rapid review. July 2022.

*This report can be downloaded here:* https://healthandcareresearchwales.org/wales-covid-19-evidence-centre-report-library

*Disclaimer:* The views expressed in this publication are those of the authors, not necessarily Health and Care Research Wales. The WCEC and authors of this work declare that they have no conflict of interest.

**TOPLINE SUMMARY:** *What is a Rapid Review?:* Our rapid reviews use a variation of the systematic review approach, abbreviating or omitting some components to generate the evidence to inform stakeholders promptly whilst maintaining attention to bias. They follow the methodological recommendations and minimum standards for conducting and reporting rapid reviews, including a structured protocol, systematic search, screening, data extraction, critical appraisal, and evidence synthesis to answer a specific question and identify key research gaps. They take 1-2 months, depending on the breadth and complexity of the research topic/ question(s), extent of the evidence base, and type of analysis required for synthesis.

*Who is this summary for?:* The Department of Health and Social Care (DHSC), who have previously created a **COVID-19 morbidities model to support the COVID-19 pandemic response**. It will also inform Welsh Government policy through work conducted by the Technical Advisory Cell.

*Background / Aim of Rapid Review:* The **COVID-19 morbidities model** has been widely used since 2020 to support Test and Trace and assess the cost-effectiveness of the COVID-19 vaccination programme. The current iteration of the Long COVID model covers several morbidities associated with COVID-19, which are essential to plan for elective care in the future and identify which services to prioritise. However, there are **uncertainties in the model around the long-term health-related quality of life (HRQoL) impact of COVID-19, which is primarily based on data for severe COVID disease or hospitalised patients at present**. The COVID-19 morbidities model requires updating to address gaps and reflect the latest HRQoL evidence. The aim of this Rapid Review was to provide updated HRQoL evidence for the COVID-19 morbidities model to better support decision-making in relation to COVID-19 policy. The latest edition of the model was published by the DHSC team in December 2020. The review focused on studies reporting on the **long-term impact on HRQoL of patients who had experienced mild symptoms or were not treated in hospital**. Inclusion was limited to studies that used validated HRQoL measures, which can be mapped onto EuroQol Quality of Life Measure – 5 dimensions (EQ-5D) and conducted in OECD countries. Two existing systematic reviews were used to identify relevant primary studies published before January 2021, with new searches focusing on the period between January 2021 to June 2022.

*Key Findings:* Thirteen primary studies were identified. Extent of the evidence base

- Most studies (n=8) were cross-sectional surveys or reported on HRQoL outcomes at a single time point post-COVID (n=2). **Only three studies (one of which was a case report) provided longitudinal follow-up data**, which included changes from baseline or reported data at multiple time points.
- **Only two studies reported on HRQoL beyond six months follow-up**: One study reported data at three months, six months, and twelve months follow-up and one study measured outcomes at six to eleven months. Five studies measured HRQoL at three months post COVID-19, one at four months, and one at five months. Three studies reported data at two months or less post COVID-19.
- **Two studies (one was a case report) focused solely on patients with mild infection**, whilst the remaining eleven studies also included patients with moderate or severe/critical COVID-19 illness. Three studies included participants categorised as non-hospitalised or hospitalised patients. twelve studies recruited patients attending outpatients or health care settings; one study recruited a general Swedish population who had a previous COVID-19 infection.
- The studies were conducted in Turkey (n=2), Denmark (n=1), Sweden (n=1), USA (n=2), Chile (n=1), Ukraine (n=1), Mexico (n=1), Austria (n=2), and The Netherlands (n=2). No UK-based studies were identified. Recency of the evidence base

- Three studies published in 2022 were conducted in 2021 (Akova & Gedikli, 2022; Bileviciute-Ljungar et al., 2022; Tanriverdi et al., 2022). Summary of results

- People who had an initial mild COVID-19 illness or were not treated in hospital can have a decreased HRQoL post-COVID. However, the extent, severity, and duration of this is not consistent. Best evidence available

- Han et al., (2022) **recruited outpatients who had mild initial COVID-19 disease** and measured **HRQoL at six to eleven months follow-up;** 436/2092 (21%) outpatients responded to the survey. The findings indicated that the burden of **persistent symptoms was significantly associated with poorer long-term health status, poorer quality of life**, and psychological distress.
- Siegerink et al., (2021) **measured HRQoL at three months, six months, and twelve months follow-up**, and recruited **patients presenting at hospital** with COVID-19, a proportion of whom were **not hospitalised**. At **three months follow-up, 22% (n=9)** of the non-hospitalised group reported **abnormal Hospital Anxiety and Depression Scale (HADS) scores** (cut-off at 16). After six months, this decreased to 16% (for n=4), and **14.8% at twelve months** (n=4).
- Labarca et al., (2021) reported a **change from baseline in percentage satisfaction with HRQoL**. They found **50% of the (n=18) ‘mild’ (non-hospitalised) COVID-19 patients reported an individual change in HRQoL**, categorised as a change of ≥ 10% on a Visual Analogue Scale (VAS) **at four months follow-up**.

*Policy Implications:*

- An initial mild COVID-19 illness can lead to a reduction in HRQoL and impaired mental health, but there is evidence indicating that patients can show significant recovery up to normal levels after one year.
- Employers should be aware that employees may have prolonged experiences of impaired mental health, including anxiety, depression, and fatigue, following COVID-19 disease, even if their initial disease was ‘mild’ (not hospitalised).
- Public health agencies should make patients with mild COVID-19 disease aware of the potential for ongoing symptoms and ways to mitigate and manage them through raised awareness and education.
- Health Boards should review their provision of long-COVID services in relation to the extent of impacts identified.
- Better quality studies that report longitudinal follow-up data on HRQoL for a representative cohort of patients who have had mild COVID-19 are required.

*Strength of Evidence:*

- The evidence on the long-term impact of a mild COVID-19 infection on HRQoL is uncertain.

## 1. BACKGROUND

COVID-19 is a coronavirus affecting individuals worldwide since the first few cases were observed in Wuhan, China in 2019 (Xiong et al., 2021). Since that time, evidence has mounted that as well as causing illness in the acute phase, some symptoms are long-lasting. There is a growing evidence-base documenting the long-term effects of COVID-19 on physical health, mental health, and quality of life (Han et al., 2022; Malik et al., 2022; Walle-Hansen et al., 2021). As part of this global evidence-base, a Rapid Evidence Summary was conducted leading to the Rapid Review question of ‘What is the long-term impact of COVID-19 on the Health-Related Quality of Life of individuals with mild symptoms (non-hospitalised)?’ This question was suggested by stakeholders from the Welsh Government and the Department of Health and Social Care (DHSC), who worked with a COVID-19 morbidities model to support the COVID-19 pandemic response. The model has been widely used since 2020 to support the Test and Trace programme and inform Welsh Government policy. In June 2022, it was recognised by the stakeholder group that the parameters of the COVID-19 morbidities model needed to be updated to address gaps and reflect more recent evidence in areas, including:

1. Health-Related Quality of Life (HRQoL) as measured by standardised outcome measures in the acute phases of COVID-19 disease
2. HRQoL of individuals hospitalised versus those with mild disease
3. HRQoL in children and young people
4. HRQoL in different ethnic groups

A Rapid Evidence Summary conducted in May 2022 showed a persistent gap in the evidence regarding HRQoL of children and young people, ethnic minority groups, and those who had mild initial COVID-19 disease. Following the stakeholder meeting (held on 1^st^ June 2022), a decision was made to focus on the long-term impact of COVID-19 on the HRQoL of those who had mild initial symptoms of the disease (not treated in hospital). Common symptoms of COVID-19 include fatigue, cough, shortness of breath, headache, anosmia, fever/chills, chest pain, rhinorrhoea, muscle aches, ageusia, sore throat, nausea/vomiting, diarrhoea and altered consciousness/confusion (Darley et al., 2021). A pre-print from 2022 suggested that 15% of individuals with mild symptoms of COVID-19 disease had not fully recovered 12 months after the initial acute infection (Hanson et al., 2022). It is estimated that 1.8 million people living in private households in the UK (2.8% of the population) were experiencing self-reported ‘long-COVID’ as of 3^rd^ April 2022 (Office for National Statistics, 2022). Long-COVID is a term used to describe signs and symptoms in adults or children that develop or persist after acute COVID-19 lasting more than four weeks after a confirmed or suspected case of COVID-19 (National Institute for Health and Care Excellence et al., 2022).

HRQoL measures are standardised validated questionnaires completed by patients to provide information on their perceived functional well-being and health status. These questionnaires can address various aspects of self-reported health, including symptoms, QoL, functionality, and physical, mental, and social well-being (Bowling, 2017). HRQoL measures play an important role in increasing patient engagement, improving health systems, and ensuring that clinical care and research are person-centred (Bray et al., 2020). Health Technology Assessment organisations, such as the National Institute for Health and Care Excellence (NICE), typically request that economic evaluations should calculate the incremental cost per quality adjusted life year (QALY) (Dakin et al., 2018; Whitehead & Ali, 2010). In order to calculate QALYs, preference-based measures are needed. In the absence of a preference-based measure, many HRQoL tools can be mapped onto the EQ-5D-5L (EuroQol Research Foundation, 2018) or the SF-6D (Brazier et al., 2002) to predict utility scores to generate QALYs (Dakin et al., 2018).

### 1.1 Purpose of this review

The aim of this Rapid Review is to provide updated evidence for the COVID-19 morbidities model regarding mild COVID-19 symptoms and HRQoL to better support decision-making in relation to COVID-19 policy.

## 2. RESULTS

### 2.1 Overview of the Evidence Base

This Rapid Review extends the findings from two previous systematic reviews (SR) published in 2021 and 2022 (Bourmistrova et al., 2022; Poudel et al., 2021). These reviews were used as the starting point for identifying relevant primary studies. Literature searches for new studies published between January 2021 and June 2022 were also conducted. Two of the included studies were found in the Bourmistrova et al. (2022) and Poudel et al. (2021) SRs (Tanriverdi et al., 2022; Van Den Borst et al., 2021).

The searches for relevant studies, and eligibility criteria used, are described in Section 5 of this report (Rapid Review methods). HRQoL measures are described in more detail in Appendix 1. Resources Searched for this Rapid Review are indicated in Appendix 2. A ‘map’ of the mild initial COVID-19 and HRQoL evidence is presented in Appendix 3. Appendix 4 presents the quality appraisals for the existing reviews and includes primary studies.

Twelve descriptive cohort studies and one case report study met the inclusion criteria (for this current Rapid Review (Akova & Gedikli, 2022; Attauabi et al., 2022; Bileviciute-Ljungar et al., 2022; Han et al., 2022; Labarca et al., 2021; Matviyets & Matiukha, 2022; Mayer et al., 2021; Ordinola Navarro et al., 2021; Rass et al., 2021, 2022; Siegerink et al., 2021; Tanriverdi et al., 2022; Van Den Borst et al., 2021). These primary studies, summarised in Table 1 and in Section 6.2, will be discussed in terms of date of data collection and the HRQoL findings.

**Table 1.**
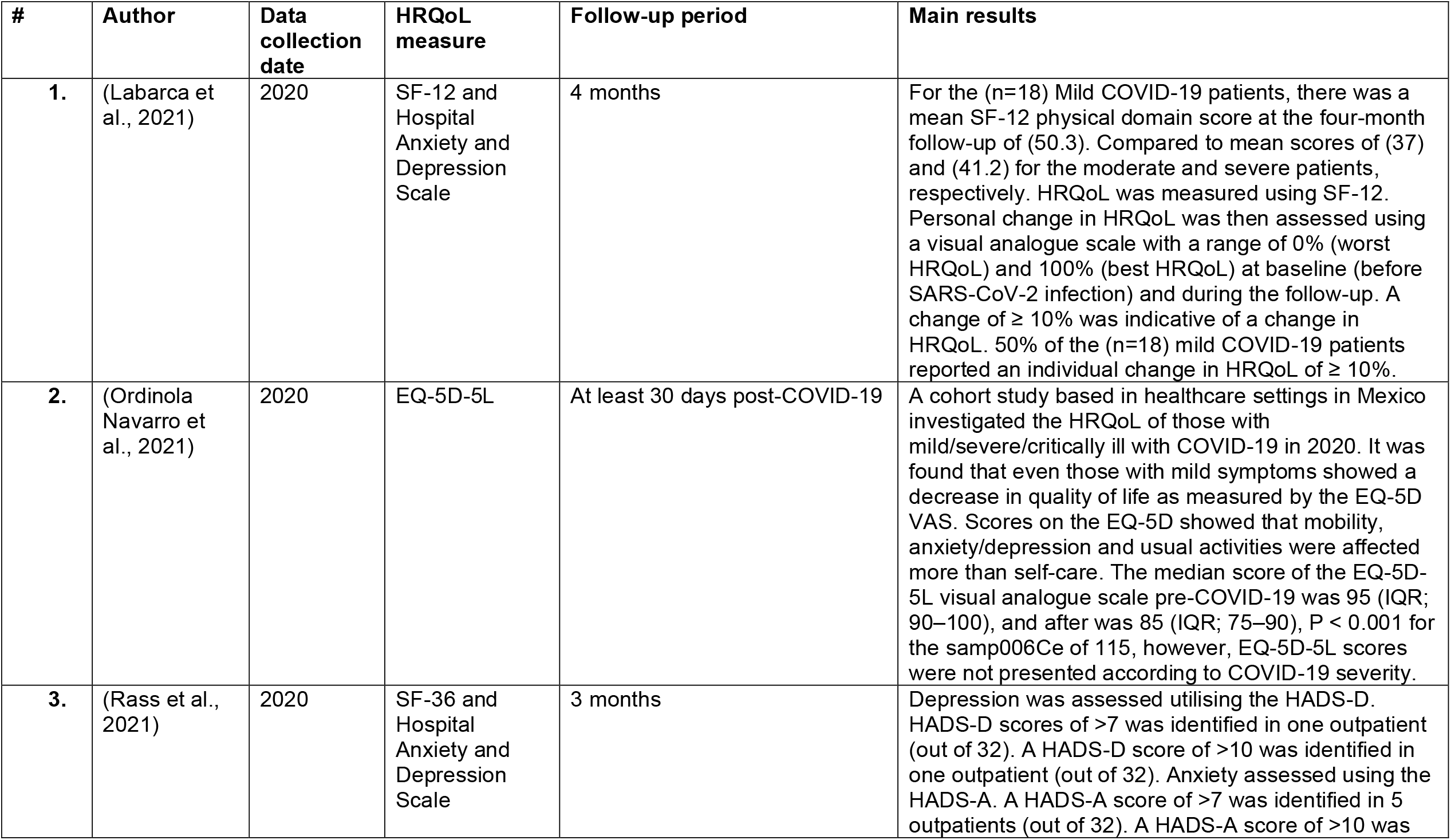
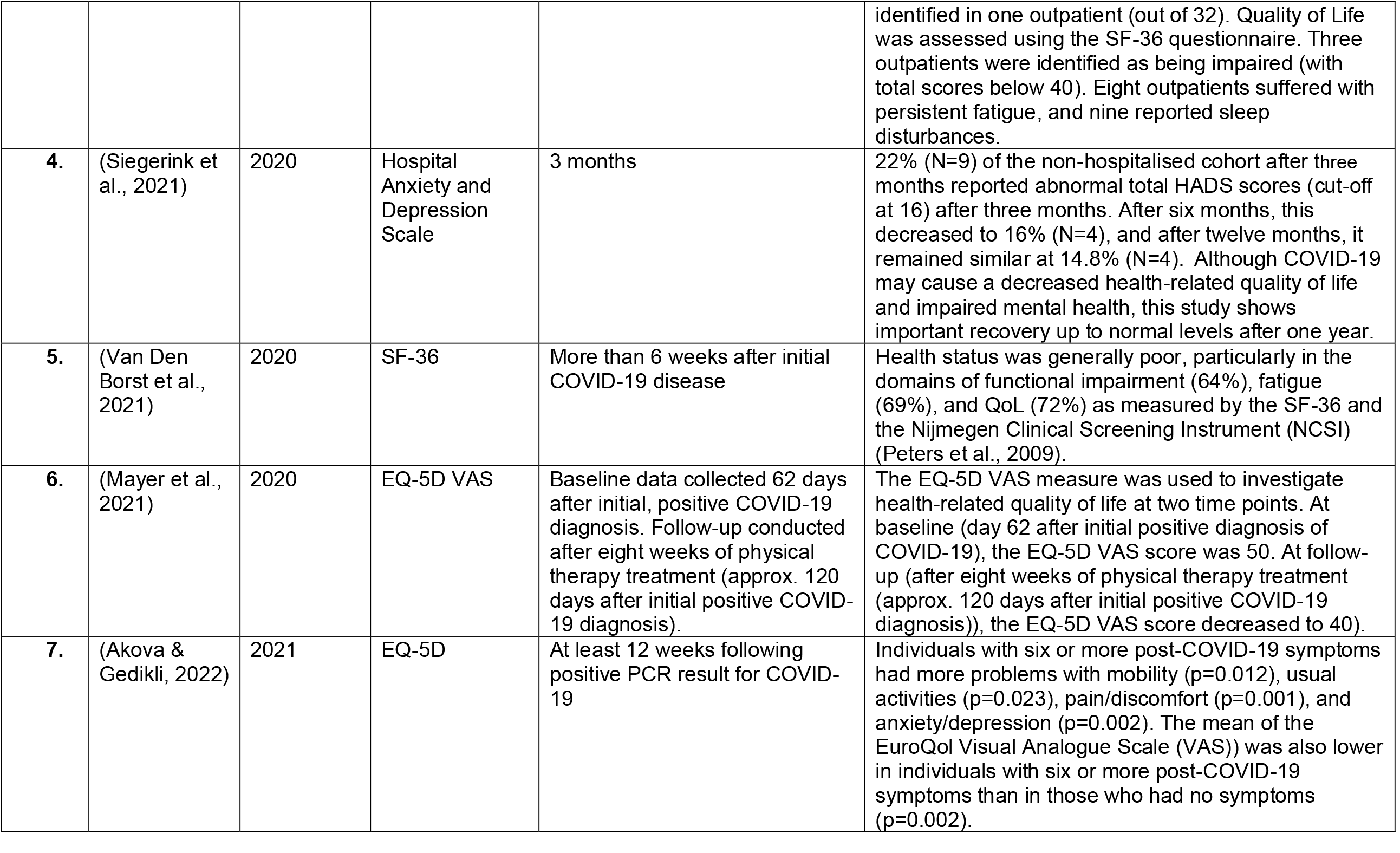
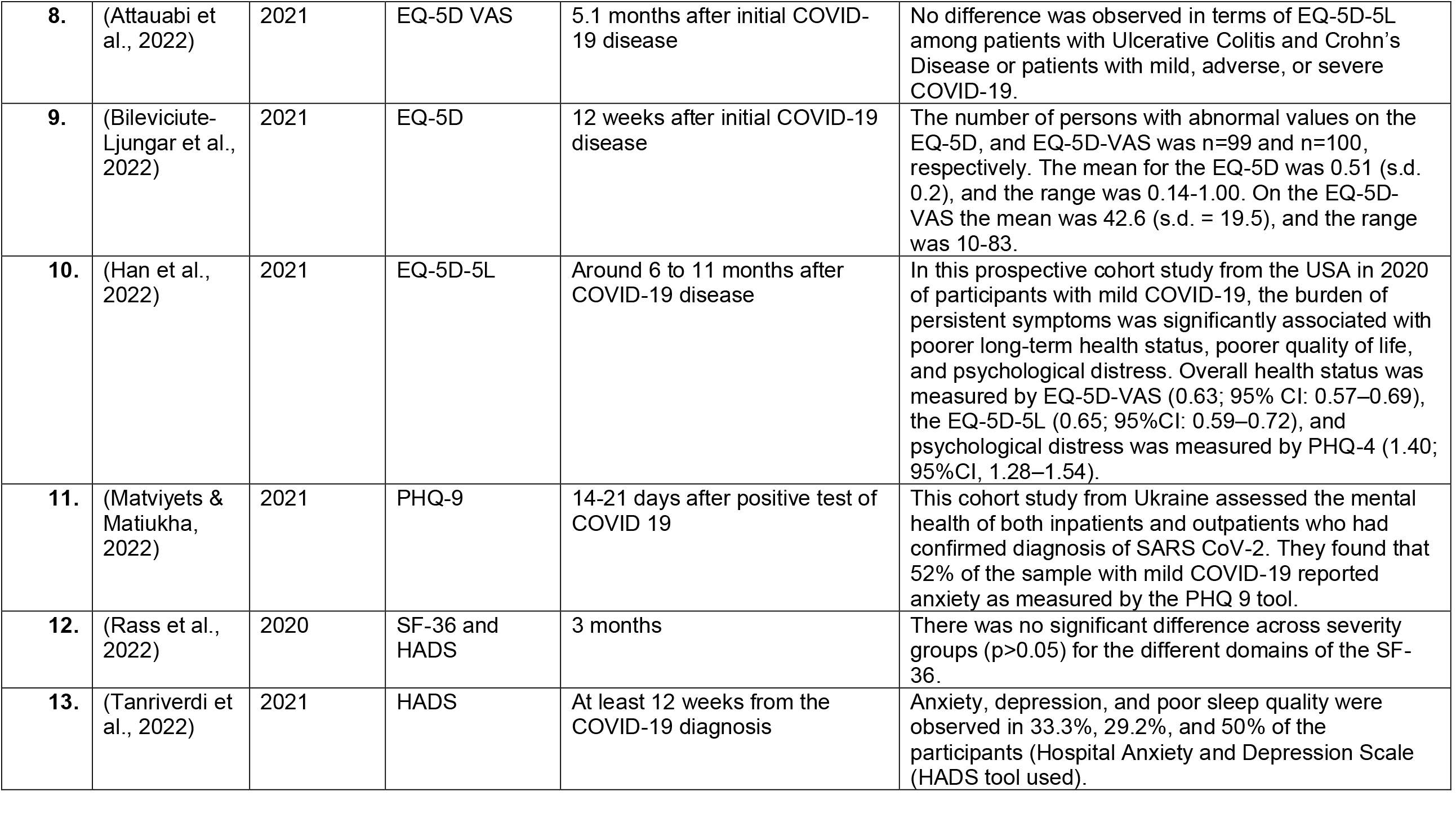
Overview of included primary papers from 2021 and 2022 including Health-Related Quality of Life measure and main results.

The studies reported in this review are grouped according to the year the study was conducted, this is sometimes different to year of publication because of the length of the publication process. Different variants of COVID-19 have been circulating since December 2019. For example, the Delta variant in 2019 was different in symptomology from the Omicron BA.5 variant circulating in the Summer of 2022. This purposeful separation of publications according to year could allow stakeholders to vary data in the model depending on the year the data was collected. The data extraction tables (see Section 6) highlight when the data was collected for each of the included studies.

#### Studies conducted in 2020

Five of the included studies reported data collected between 2019 and 2020 (González et al., 2021; Ordinola Navarro et al., 2021; Rass et al., 2021; Siegerink et al., 2021; Van Den Borst et al., 2021). One of the included studies was a case report (Mayer et al., 2021). These cohort studies reported on HRQoL measured by a variety of outcome measures, including EQ-5D, SF-36 and the Hospital Anxiety and Depression Scale (HADS), all of which could be mapped against the EQ-5D with individual-level data. The HRQoL results of the studies are reported below according to the measure used.

#### EQ-5D

A cohort study based in healthcare settings in Mexico investigated the HRQoL of patients (n=115) who were categorised as being mild, severe and critically ill with COVID-19 in 2020 (Ordinola Navarro et al., 2021). It was found that even those with mild symptoms showed a decrease in QoL as measured by the EQ-5D Visual Analogue Scale (VAS). Scores on the main EQ-5D-5L measure showed that mobility, anxiety, depression, and usual activities were affected more than self-care. The median score of the EQ-5D-5L visual analogue scale pre-COVID-19 was 95 (IQR; 90–100), and after was 85 (IQR; 75–90), P < 0.001 for the sample of 115, however, EQ-5D-5L scores were not presented according to COVID-19 severity.

A prospective cohort study was conducted in the USA in 2020 (Han et al., 2022) six to eleven months after COVID-19 disease. This study investigated the HRQoL of 213 outpatients who had mild initial COVID-19 disease. The authors measured HRQoL status using the EQ-5D-VAS (mean score = 0.63; confidence interval 95%: 0.57–0.69), and the EQ-5D-5L (mean score: 0.65; confidence interval 95%: 0.59–0.72) around 6 to 11 months after acute infection.

A case report of a female who had mild initial COVID-19 in the USA found that her HRQoL scores measured by EQ-5D VAS and PTSD scores did not improve following eight weeks of physiotherapy (Mayer et al., 2021). This was despite improvements in physical functioning. This case report was published in 2021, but the dates of data collection were unclear.

#### Hospital Anxiety and Depression Scale (HADS)

A prospective observational cohort study was conducted during the first wave of COVID-19 in 2020 in The Netherlands with patients who presented at hospital with COVID-19 but were not hospitalised (Siegerink et al., 2021). HRQoL was measured using the HADS questionnaire. At three months follow-up, 22% (n=9) of the non-hospitalised cohort reported abnormal total HADS scores (cut-off at 16). After 6 months, this decreased to 16% (n=4), and after 12 months fell further to 14.8% (n=4). The authors noted that although COVID-19 correlated with decreased HRQoL and impaired mental health, this study provided evidence that individuals recovered to pre-acute COVID-19 levels after one year.

#### SF-12 and SF-36 outcome measures

A study conducted in the Netherlands in 2020 investigated the health and QoL status of patients (n=124) hospitalised and not hospitalised for COVID-19 (Van Den Borst et al., 2021). Twenty-seven patients had mild initial COVID-19. For the group as a whole, the six weeks post COVID-19 health status was generally poor, particularly in the domains of functional impairment (64%), fatigue (69%), and QoL (72%) as measured using the SF-36 and the Nijmegen Clinical Screening Instrument (NCSI) (Peters et al., 2009). Although some sub-group analyses were conducted, these were not clearly presented in terms of mild, moderate, and severe cases. Patients with mild disease referred by GPs to hospital were younger than patients with moderate-to-critical disease and were predominantly female. The authors noted that longer follow-up studies are warranted to explain pathways and to find predictors of complicated long-term trajectories of recovery.

An Austrian study conducted in 2020 included participants (n=135) with COVID-19 recruited three months after presenting at hospital (Rass et al., 2021). A proportion of these participants (n=32; 24%) were treated as outpatients and thus classified as non-hospitalised. At the three-months follow-up, depression was assessed using the HADS-D measure. A HADS-D score of >7 (0-7 = ‘normal’) was identified in one outpatient (out of 32). A HADS-D score of >10 (8-10 = ‘borderline abnormal’ and 11-21 = ‘abnormal) was identified in one other outpatient (out of 32). Anxiety was assessed using the HADS-A measure. Five outpatients scored 8-10 on this measure (indicating mild anxiety), and one outpatient gave a score over 10 (indicating a clinically meaningful anxiety disorder). QoL was assessed using the SF-36 questionnaire. Three outpatients were identified as being impaired (with total SF-36 scores below 40).

The SF-12 and the HADS measures were used in a study conducted in hospital and community settings in Chile with 60 patients from April to June 2020 (Labarca et al., 2021), four months following initial COVID-19 disease. Of the 60 patients, 18 had mild COVID-19 disease, 17 had moderate COVID-19 disease, and 25 had severe COVID-19 disease. The authors found that patients diagnosed with COVID-19 presented a high prevalence of symptoms and affected anxiety and depression status (as measured by the HADS) regardless of initial infection severity. For the (n=18) mild COVID-19 patients, there was a mean SF-12 physical domain score at the four-month follow-up of (50.3). Compared to mean scores of (37) and (41.2) for the moderate and severe patients, respectively, with a higher score reflecting a better quality of life. A Visual Analogue Scale (VAS) was used to assess personal change in HRQoL, with a range of 0% (worst HRQoL) and 100% (best HRQoL). Fifty percent of the patients with initial mild COVID-19 disease (n=18) reported an individual change in HRQoL. A score change of ≥ 10% was indicative of a change in HRQoL as measured by the SF-12.

#### Studies conducted in 2021

Seven of the included primary papers were cohort studies reporting data collected in 2021 or published in 2022, where the date of data collection was not reported (Akova & Gedikli, 2022; Attauabi et al., 2022; Bileviciute-Ljungar et al., 2022; Han et al., 2022; Matviyets & Matiukha, 2022; Rass et al., 2022; Tanriverdi et al., 2022). The EQ-5D measure was used in three of the studies described below.

#### EQ-5D

A study from Turkey conducted in March 2021 included 151 adults from at least 12 weeks following positive PCR result for COVID-19 (and not treated in hospital) (Akova & Gedikli, 2022). The authors found that individuals with six or more post-COVID-19 symptoms had more problems with mobility (p=0.012), usual activities (p=0.023), pain/discomfort (p=0.001), and anxiety/depression (p=0.002) compared to individuals with fewer post-COVID-19 symptoms as measured by EQ-5D (Akova & Gedikli, 2022). The mean score was also lower in individuals with six or more post-COVID-19 symptoms than in those who had no symptoms (p=0.002). The authors noted that patient follow-up should be given importance, especially for females and the elderly, as the more symptoms that persist, the lower the HRQoL.

A survey study conducted in Sweden between April and August 2021 with 100 patients with mild illness found that 99 patients reported abnormal values on the EQ-5D and 100 patients reported abnormal values on the EQ-5D-VAS (Bileviciute-Ljungar et al., 2022) 12 weeks after acute infection. The mean for EQ-5D was 0.51 (s.d. = 0.2), and the range was 0.14-1.00. On the EQ-5D-VAS, the mean was 42.6 (s.d. = 19.5), and the range was 10-83. Population norms in Sweden for the EQ-5D is 0.85, and the EQ-5D-VAS is 85 for people between 35 and 54 years old (Szende et al., 2014). Pain was often reported by post-COVID-19 sufferers despite a mild initial infection.

A study from Denmark (Attauabi et al., 2022) investigating n=222 patients with inflammatory bowel symptoms, including Ulcerative Colitis (UC) and Crohn’s Disease (CD), found no difference in HRQoL as measured by the EQ-5D-5L among patients with UC and CD or patients with mild, adverse, or severe COVID-19, after 5.1 months. The major limitation of this study is that those with pain were perhaps more inclined to complete a questionnaire on the topic.

#### Hospital Anxiety and Depression Scale (HADS)

A Turkish study conducted in February 2021 with a small sample of adults (n=25) who had mild initial COVID-19 diagnosis (Tanriverdi et al., 2022) found that anxiety, depression, and poor sleep quality were observed in 33.3%, 29.2%, and 50% of the participants as measured by the HADS and Pittsburgh Sleep Quality Index, respectively, at least 12 weeks from COVID-19 diagnosis. The authors found that poor sleep quality was the main negative consequence of having had a past COVID-19 diagnosis.

### 2.2 Bottom line results for HRQoL and mild initial COVID-19

**Studies relating to HRQoL and mild initial COVID-19 were published in 2021 and 2022 based on data from 2020 to June 2022. Many studies found that mild COVID-19 was associated with decreased quality of life, and some studies showed no difference in quality of life between those with severe and mild COVID-19. Notably, some of the studies included in this Rapid Review showed that patients with previous symptoms of COVID-19 improved in terms of HRQoL over time post initial infection**.

## 3. DISCUSSION

### 3.1 Summary of the findings

This RR was built on the foundations of the Systematic Reviews by Bourmistrova et al. (2022) and Poudel et al. (2021).

In this review, thirteen of the included studies involved individuals who had mild initial COVID-19 disease (or not treated in hospital) and whose quality of life was measured by a validated HRQoL outcome measure. The cohort studies such as Labarca et al (2021) suggest that mild COVID-19 infection may influence future health.

### 3.2 Limitations of the available evidence

All of the included evidence was collected during different time points of the COVID-19 pandemic (between January 2020 and June 2022), when different variants were prevalent in different countries worldwide. As such, this research provides an overview of the HRQoL of those patients with mild forms of the different variants. Therefore, for example, the authors cannot be certain that the mild form of Omicron was different to the mild form of Delta in terms of recovery after mild COVID-19 disease. Moreover, the vaccination programmes did not start simultaneously in every country; therefore, some earlier studies would include unvaccinated cohorts, and some later studies may include mostly or partly vaccinated cohorts. Also, there has been some variance in the administration of the rollout of vaccination programmes worldwide.

This review found very limited evidence on the long-term impact of mild COVID-19 and HRQoL. Only three studies (one of which was a case report: Mayer et al., 2021) provided longitudinal follow-up data, which included change from baseline (Labarca et al., 2021; Mayer et al., 2021) or data on multiple time points (Siegerink et al., 2021). The other included studies provided some evidence (which could provide a utility at this point in time) as there was a lack of longitudinal studies with repeated measures. This kind of cross-sectional evidence was included because comparisons could be made to general population value sets. Few studies limited inclusion to people with mild COVID-19, with many also including patients with severe illness or hospitalised patients, with outcome data for specific sub-populations being inadequately reported. Most studies recruited patients with a previous mild illness attending outpatient clinics or used postal questionnaires, which will likely result in a higher proportion of patients with adverse impact of COVID-19 being included.

There was limited information regarding how the mild initial COVID-19 illness affected different population groups. Although most studies focused on adults, age and ethnicity were not the focus of most of the studies. Therefore, sub-group analyses addressing HRQoL effects of mild COVID-19 in members of minority groups are not available.

### 3.3 Implications for policy and practice

- An initial mild COVID-19 infection can lead to a decrease in HRQoL and impaired mental health, but there is evidence indicating that patients can show important recovery up to normal levels after one year.
- Employers should be aware that employees may have prolonged experiences of impaired mental health, including anxiety, depression and fatigue following COVID-19 disease, even if their initial disease was of mild severity.
- Public health agencies should make patients with mild COVID-19 disease aware of the potential for ongoing symptoms and ways to mitigate and manage them through raised awareness and education.
- Health Boards should review their provision of long-COVID services in relation to the extent of impacts identified.

### 3.4 Strengths and limitations of this Rapid Review

#### Strength of the rapid review

- This Rapid Review was conducted in June 2022 and covers the first waves of the COVID-19 pandemic, including data from January 2020 to June 2022.
- The focus of this Rapid Review was the long-term HRQoL of patients who tested positive for COVID-19 and experienced mild symptoms. All studies included reported validated HRQoL measures (see Appendix 1) that can be mapped onto EQ-5D.
- All the included studies were from peer-reviewed journals.

#### Limitation of the rapid review

- An important limitation of this review is the lack of available studies reporting longitudinal follow-up data for patients who have had a mild initial illness or not treated in hospital.
- There is an acknowledgement that the availability of studies including individuals with mild initial COVID-19 could restrict the number of studies available to include in a Rapid Review.
- People with mild initial COVID-19 disease may not have sought treatment or attended any healthcare services due to the nature of their illness and being asked to stay at home. They may also have chosen not to test for COVID-19, and in the first wave of COVID-19 there were no readily available methods of testing for a positive diagnosis. Therefore, it would be difficult to identify and recruit patients with mild infection outside of healthcare settings, with risk of selection bias in study recruitment.
- This Rapid Review only included studies from Organisation for Economic Co-operation and Development (OECD) countries. However, data were also available from, for example, Brazil (Barreto et al., 2021), China (Chen et al., 2022), India (Maheshwari et al., 2021) and Romania (Giurgi-Oncu et al., 2021). These may also have been of some relevance but were excluded due to being outside OECD countries. It is possible that these studies may report good quality data that may be of interest to researchers in the future.
- The rapid review used existing systematic reviews to identify studies published before January 2021. This enabled us to complete the review in a timely way and avoid duplication and research waste. However, one of the existing reviews used to identify early studies (Bourmistrova et al., 2022) focused on evaluating the mental health impact of COVID-19 and did not cover other important HRQoL domains such as physical function. This means that some relevant early studies may have been missed, although we do not believe this to be the case. We were also more interested in studies published between 2021-2022 because the current DHSC COVID-19 Morbidities model is based on data up to December 2020.

## Data Availability

All data produced in the present study are available upon reasonable request to the authors

## Abbreviations

Acronym: Full Description
CD: Crohn’s Disease
COVID-19: Coronavirus disease-19
EQ-5D: EuroQol Quality of Life Measure – 5 dimensions
GP: General Practitioner
HADS: Hospital Anxiety and Depression Scale
HADS-A: Hospital Anxiety and Depression Scale - Anxiety
HADS-D: Hospital Anxiety and Depression Scale - Depression
HRQoL: Health-Related Quality of Life
IBS: Inflammatory Bowel Diseases
Mild COVID-19: COVID-19 disease without hospitalisation (non-severe COVID-19)
NHS: National Health Service
OECD: Organisation for Economic Co-operation and Development
ONS: Office for National Statistics
PTSD: Post-Traumatic Stress Disorder
PHQ-9: Patient Health Questionnaire-9
QoL: Quality of life
SD: Standard deviation (from the mean)
SF-12: Short Form-12
SF-36: Short Form-36
UC: Ulcerative Colitis
VAS: Visual Analogue Scale
WCEC: Wales COVID-19 Evidence Centre

## 5. RAPID REVIEW METHODS

### 5.1 Eligibility criteria

The eligibility criteria for the review is presented in Table 2 and is based on the Population, Intervention, Comparison and Outcome (PICO) framework (Schardt et al., 2007). The stakeholders for this rapid review were interested in the long-term HRQoL outcomes for individuals with mild COVID-19 disease or not treated in hospital. The HRQoL scores were intended for use as part of the modelling work and to develop health utility scores to calculate QALYs. The outcomes of interest here, therefore, included the use of standardised HRQoL measures that can be used to develop a health utility score, including but not limited to EQ-5D (EuroQol Research Foundation, 2018) and SF-36 (Ware, 2000). A full list of HRQoL measures that can be mapped to EQ-5D can be found in Dakin et al., (2018) and Whitehead and Ali (2010) (Dakin et al., 2018; Whitehead & Ali, 2010).

**Table 2:** Eligibility Criteria.

| **Review question:** |  |  |
| --- | --- | --- |
| What is the long-term impact of COVID-19 on the Health-Related Quality of Life of individuals with mild symptoms (non-hospitalised)? |  |  |
|  | **Inclusion criteria** | **Exclusion criteria** |
| **Participants** | Individuals with or who have had mild COVID-19 disease (suspected or confirmed)<br>Individuals with or who have had COVID-19 disease that were not treated in hospital | Individuals who were hospitalised due to COVID-19 disease<br>Individuals who have not been infected by SARS-CoV-2 and suffered no symptoms of the disease |
| **Intervention / exposure** | Cohort of patients who have had COVID-19 disease | Individuals who have not had COVID-19 disease. |
| **Comparison** | Studies investigating HRQoL | Studies not investigating HRQoL |
| **Outcome measures** | Any Health-Related Quality of Life (HRQoL) outcome measure that could be converted into a health utility score (e.g. EQ-5D) (Dakin et al., 2018; Whitehead & Ali, 2010) | Any measure for which there is no recognised method for converting the score into a health utility (either directly, such as EQ-5D etc; or indirectly, such as via mapping) |
| **Study design** | Longitudinal studies or cross-sectional studies reporting data beyond two weeks following acute disease. |  |
| **Countries** | OECD | Non-OECD countries |
| **Other factors** | Update the Bourmistrova et al (2022) and Poudel et al (2021) SRs. Searched for new papers from January 2021. (The rapid review was intended to feed into the current morbidity model developed in December 2020 by DHSC,) | None |

Initially, this review only intended to include longitudinal studies; however, due to a lack of studies reporting longitudinal data with repeated measures, inclusion criteria were updated to include cross-sectional studies reporting data beyond two weeks following acute disease.

### 5.2 Literature search

#### 5.2.1 Incorporating studies from existing reviews

Two published systematic reviews conducted in 2021 were used as the starting point for the literature search (Bourmistrova et al., 2022; Poudel et al., 2021). Studies included in the two existing reviews were screened for inclusion in this Rapid Review. New (database) searches were then conducted to identify primary studies published from January 2021 through to June 2022

#### 5.2.2 Evidence sources

Key evidence sources include:

1. ASSIA
2. CINAHL
3. Cochrane Library
4. EmBASE
5. Medline
6. PsycInfo

Key sources were searched for papers published between 1^st^ January 2021 and 13th June 2022. The searches were limited to published research in the English or Welsh languages. The scope outlined for this search is to keep the review concise and deliverable within the timeframe expected for a Rapid Review.

#### 5.2.3 Search strategy

The search strategy for the Rapid Review is outlined below. This search strategy was created for Medline and adapted for the other databases.

##### Search strategy for Medline

1. Patients/
2. (patient*).ti,ab
3. (case* or sufferer* or victim* or infect*).ti,ab
4. 1 OR 2 OR 3
5. ((non or no*) adj2 (hospital* or admission or admittance or admitted or admit*)).ti,ab,kw
6. (mild or moderate or non-serious or untreated or non-treated).ti,ab,kw
7. 5 or 6
8. COVID-19/
9. (COVID).ti,ab
10. (coronavirus* or coronovirus* or coronaviri* or 2019-nCoV or 2019nCoV or nCoV2019 or nCoV-2019 or covid-19* or covid19* or ncov* or n-cov* or HCoV* or SARS-CoV-2 or SARSCoV-2 or SARSCov2 or SARS-CoV2 or severe acute respiratory syndrome).ti,ab
11. 8 or 9 or 10
12. Quality of life/
13. ((health or health related) adj2 (quality or quality of life)).ti,ab,kw
14. 12 or 13
15. 4 and 7 and 11 and 14

#### 5.2.4 Reference management

The Covidence systematic review software was used to store and manage citations. Duplicates were removed in Covidence (Veritas Health Innovation, 2021).

The citations were screened on title and abstract by three members of the core review team. Full-text articles were then retrieved and further assessed for inclusion or exclusion. Any queries regarding inclusion/exclusion were resolved by discussion between members of the review team.

### 5.3 Data extraction

The data were extracted from the included studies using a pre-defined data extraction tool developed to capture all relevant data (see section 6.2). Extracted data included study details such as author, year, setting, aim, design, population, and sample size. The data extraction included data specific to the review question, type of study, method of analysis, key findings, and author conclusions.

Included papers were distributed among the core review team for data extraction. A sample of extracted studies was checked against the papers for accuracy by the review lead. A proportion of the papers (10%) were double extracted to check for discrepancies between reviewers.

### 5.4 Assessment of methodological quality

Quality appraisal was carried out by members of the core review team using the JBI critical appraisal tools. This includes the JBI systematic reviews and research syntheses checklist (Joanna Briggs Institute, 2017), JBI case reports checklist (Munn et al., 2021), and JBI cohort studies checklist (Joanna Briggs Institute, 2021).

Members of the review team choose the most appropriate JBI critical appraisal tool. A quarter of critical appraisals were checked by a second reviewer. Discrepancies arising during the critical appraisal process were discussed until the review team reached an agreement.

### 5.5 Synthesis

Study characteristics and results are presented in Section 6.2, and a short narrative synthesis was developed to bring the evidence together (Mishler, 1995). Quantitative data from the included studies have been summarised and presented in the results section.

### 5.6 Assessment of body of evidence

All included primary papers were from peer reviewed journals.

## 6. EVIDENCE

### 6.1 Study selection flow chart

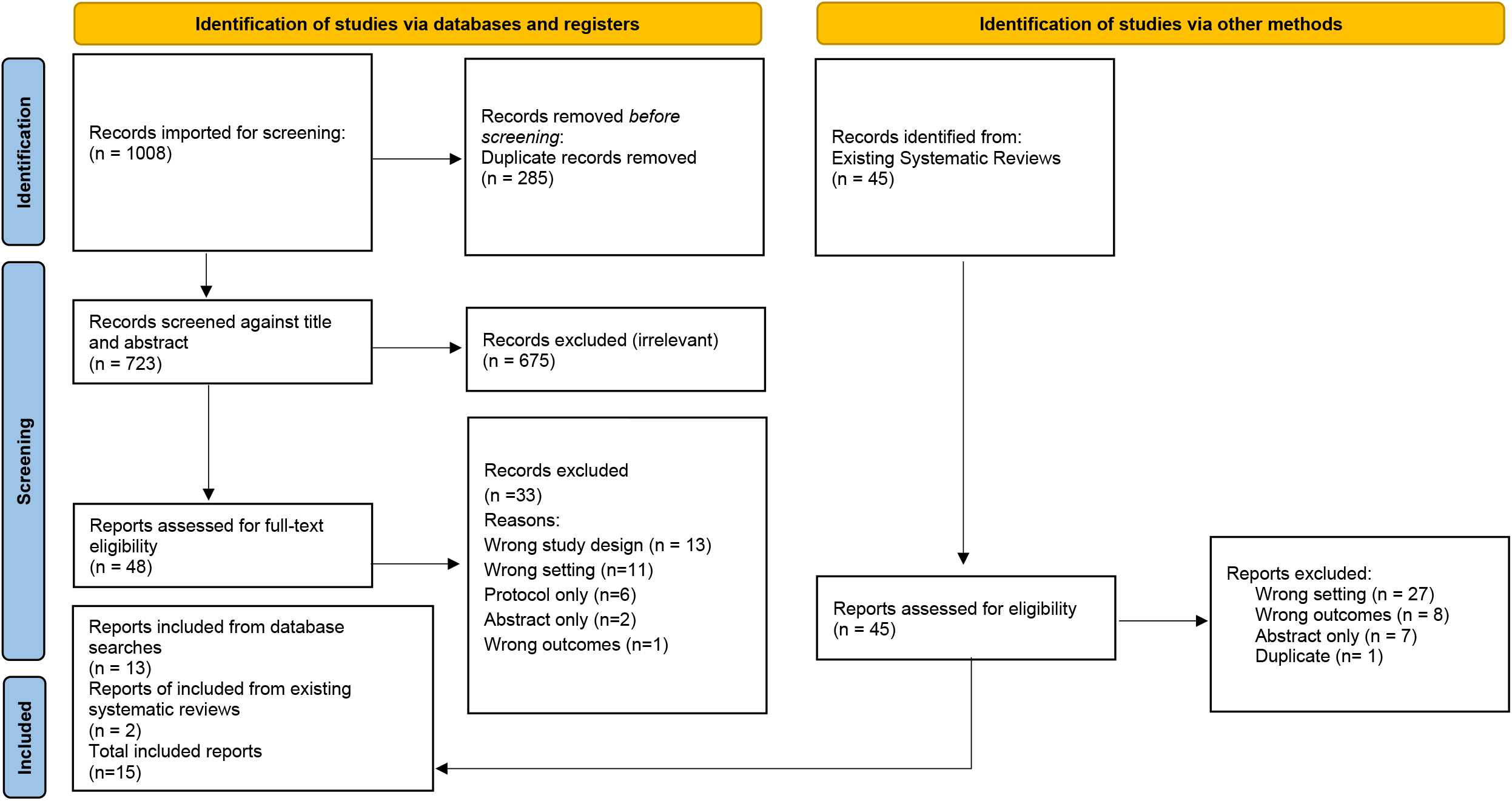

The study selection flow chart is shown as a PRISMA flow chart (Page et al., 2021).

*From:* Page MJ, McKenzie JE, Bossuyt PM, Boutron I, Hoffmann TC, Mulrow CD, et al. The PRISMA 2020 statement: an updated guideline for reporting systematic reviews. BMJ 2021;372:n71. doi: 10.1136/bmj.n71. For more information, visit: http://www.prisma-statement.org/

### 6.2 Data extraction tables

See Appendix 5 for the quality appraisal tables.

**Table A.6.1.** Secondary research – data extraction

| Citation<br>(Country) | Review details | Included studies | Quality | Findings and observations/notes |
| --- | --- | --- | --- | --- |
| Bourmistrova et al 2022<br>(UK)<br>(Bourmistrova et al., 2022) | **Review period:** The search covered studies published between 1st October 2019 and 29th August 2021.<br>**Review purpose:** Systematic review of long-term effects of COVID-19 on mental health, focussing on mild disease.<br>The long-term effect of direct COVID-19 infection has been associated with no or mild symptoms.<br>Included studies exhibited the long-term prevalence of anxiety, depression, PTSD and sleep disturbance levels.<br>**Included study designs:**<br>Retrospective cohort studies (n=2)<br>Cohort studies (n=20)<br>Controlled cohort study (n=1)<br>Cross-sectional cohort studies (n=5)<br>Case control studies (n=3)<br>Case series (n=2)<br>**Included outcome measures:**<br>HRQoL | **Number of included studies:** 33 were included in the review involving a total of 6743 participants.<br>**Key characteristics:** Hospitalised and non-hospitalised, severe and mild COVID-19 patients with regards to symptoms and quality of life. | Moderate | The overall effect of the pandemic has been associated with worsening psychiatric symptoms. However, the long-term effect of direct COVID-19 infection has been linked with no or mild symptoms. Studies exhibited the long-term prevalence of anxiety, depression, PTSD, and sleep disturbances to be comparable to general population levels. |
|  | PTSD<br>Symptoms of COVID-19 disease |  |  |  |
| **Poudel et al (2021)**<br>(England, UK)<br>(Poudel et al., 2021) | **Review period:** The authors searched for original studies published between December 2019 and Jan 2021.<br>**Review purpose:**<br>To investigate the impact on health-related quality of life (HRQoL) of patients with Covid-19.<br>**Included study designs:**<br>Observational (e.g. cross-sectional surveys)<br>Experimental study<br>**Included outcome measures:**<br>The majority of the studies (n = 10) used generic HRQoL assessment tool (five used SF-36, five EQ-5D-5L), and the rest used a pulmonary disease-specific HRQoL tool, i.e. SGRQ (St George's Respiratory Questionnaire) tool (2/12). Clinical COPD Questionnaire (CCQ) (1/12), and PROMIS tool (1/12) (2 out of the 12 studies used two HRQoL assessment tools i.e. SGRQ and EQ-5D-5L, and CCQ and EQ-5D-5L). | **Number of included studies:** 12 included papers<br>**Key characteristics:** HRQoL measures | Moderate | The health-related quality of life scores were considerably lower among the severe patients who were admitted to ICU compared to moderate patients who were admitted to general wards; the HRQoL scores were found to be lower in female Covid-19 patients and patients who were of old age.<br>This review by Poudel et al (2021) also included three papers with clinically suspected Covid-19 patients and both confirmed and suspected patients, as during the start of the Covid-19 pandemic, testing facilities were not widely available, and researchers had to include suspected Covid-19 patients as well in their research studies. |

**Table A.6.2.**
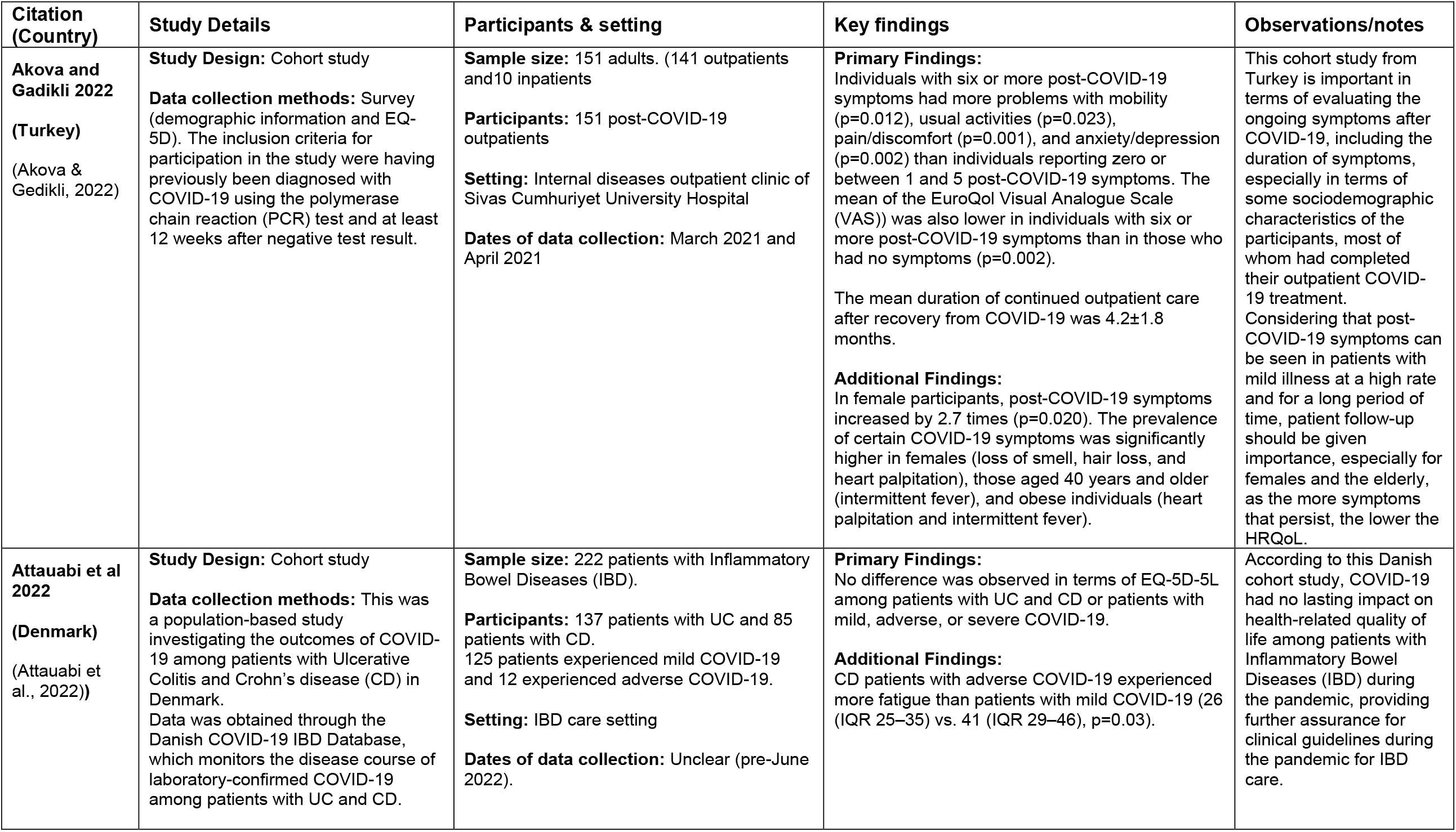
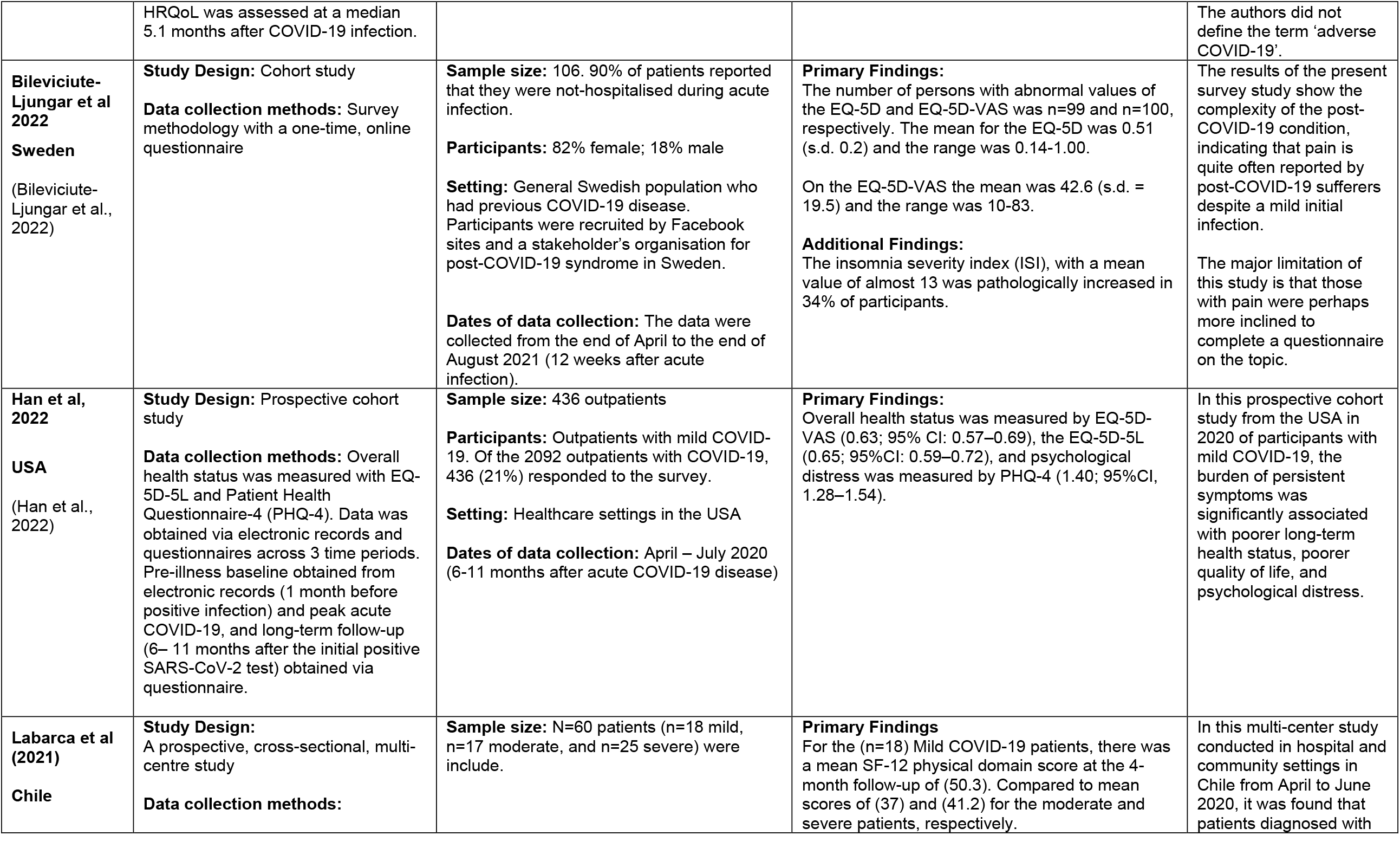
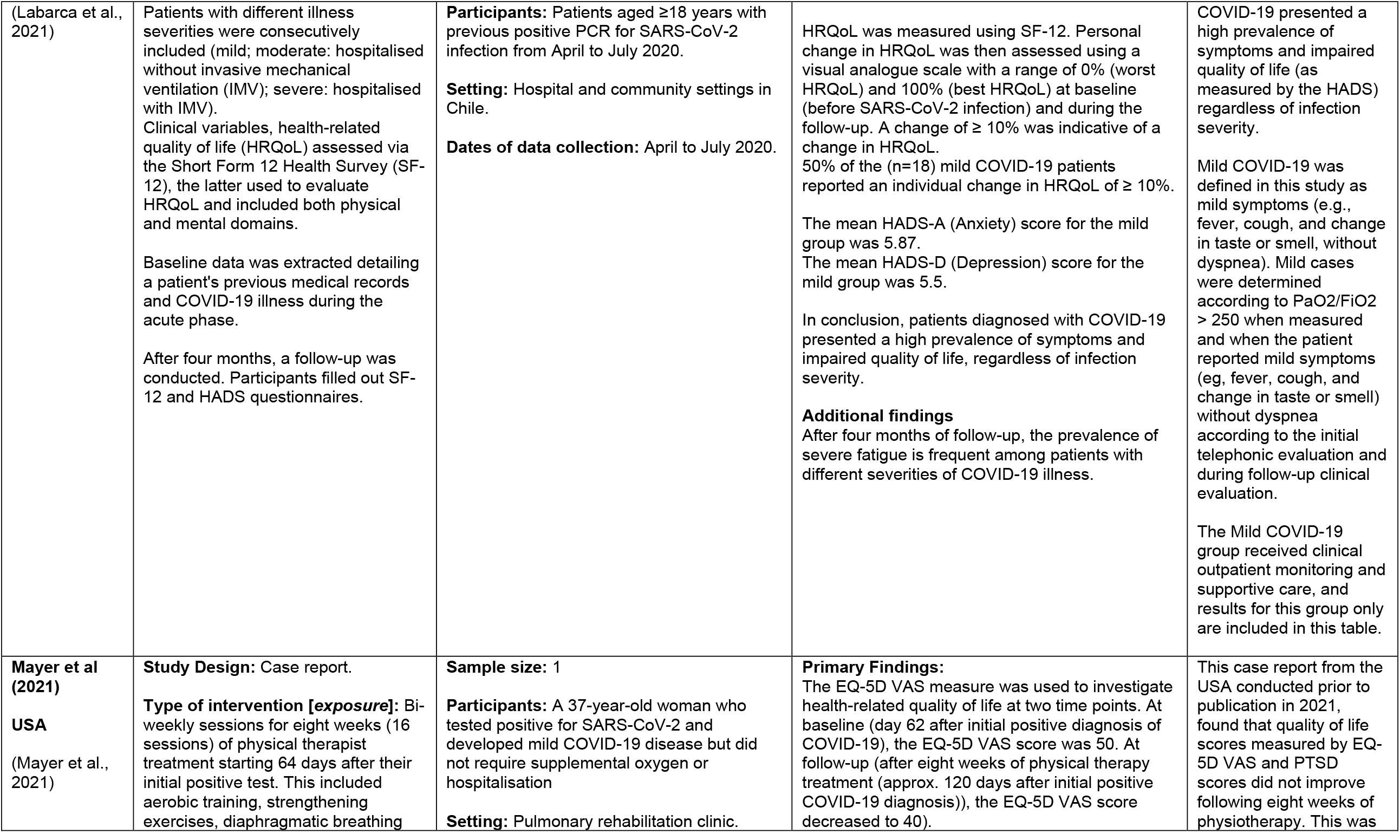
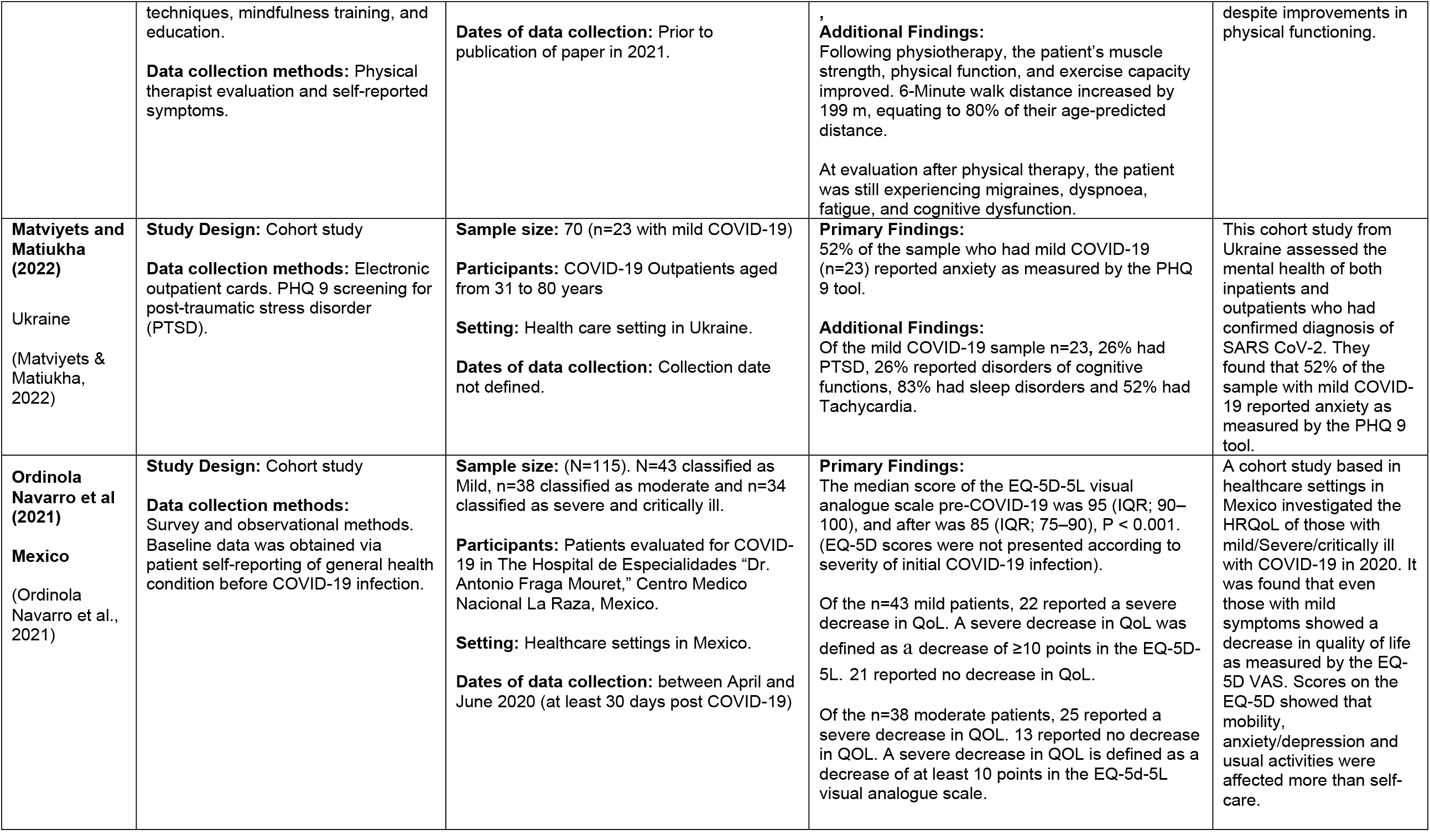
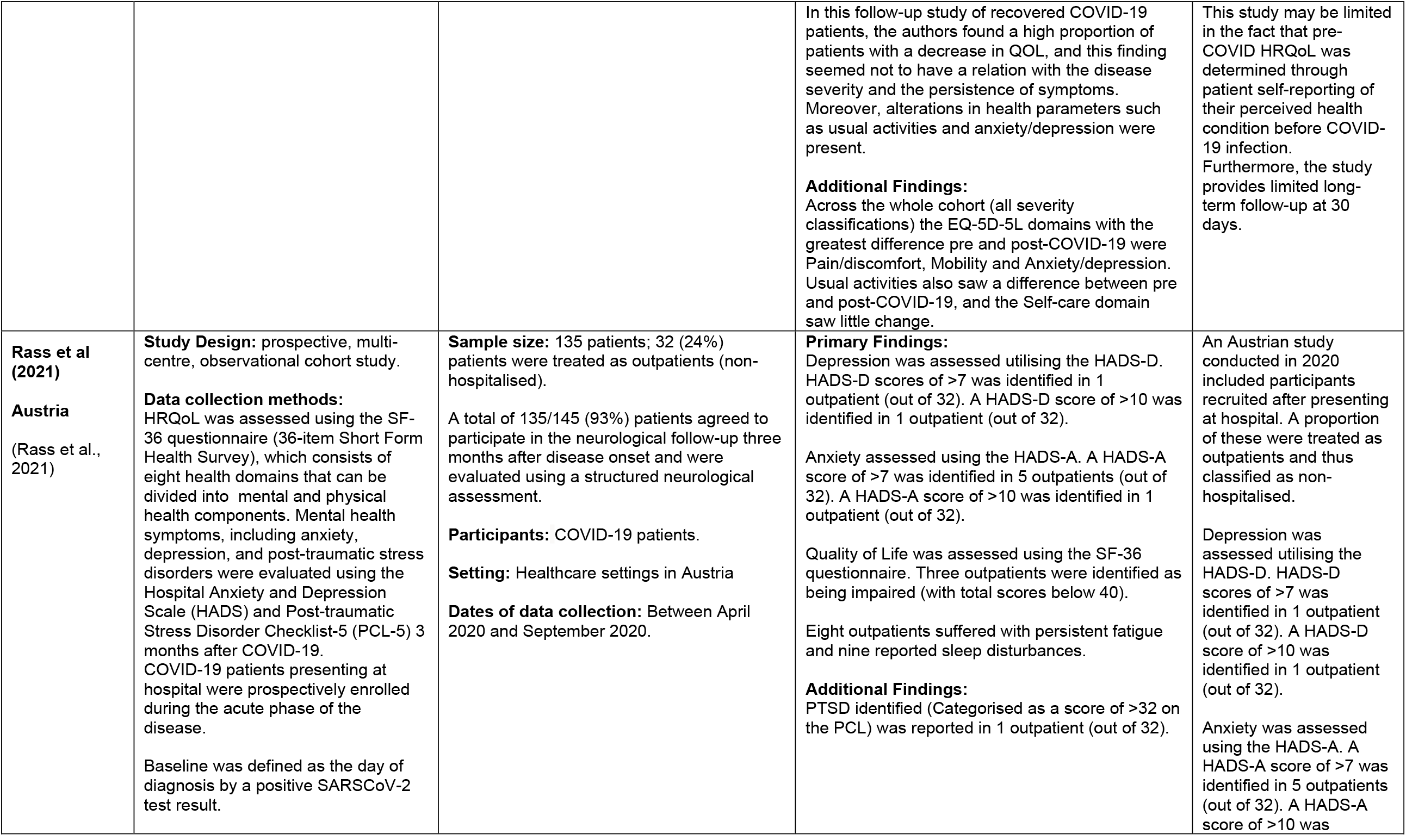
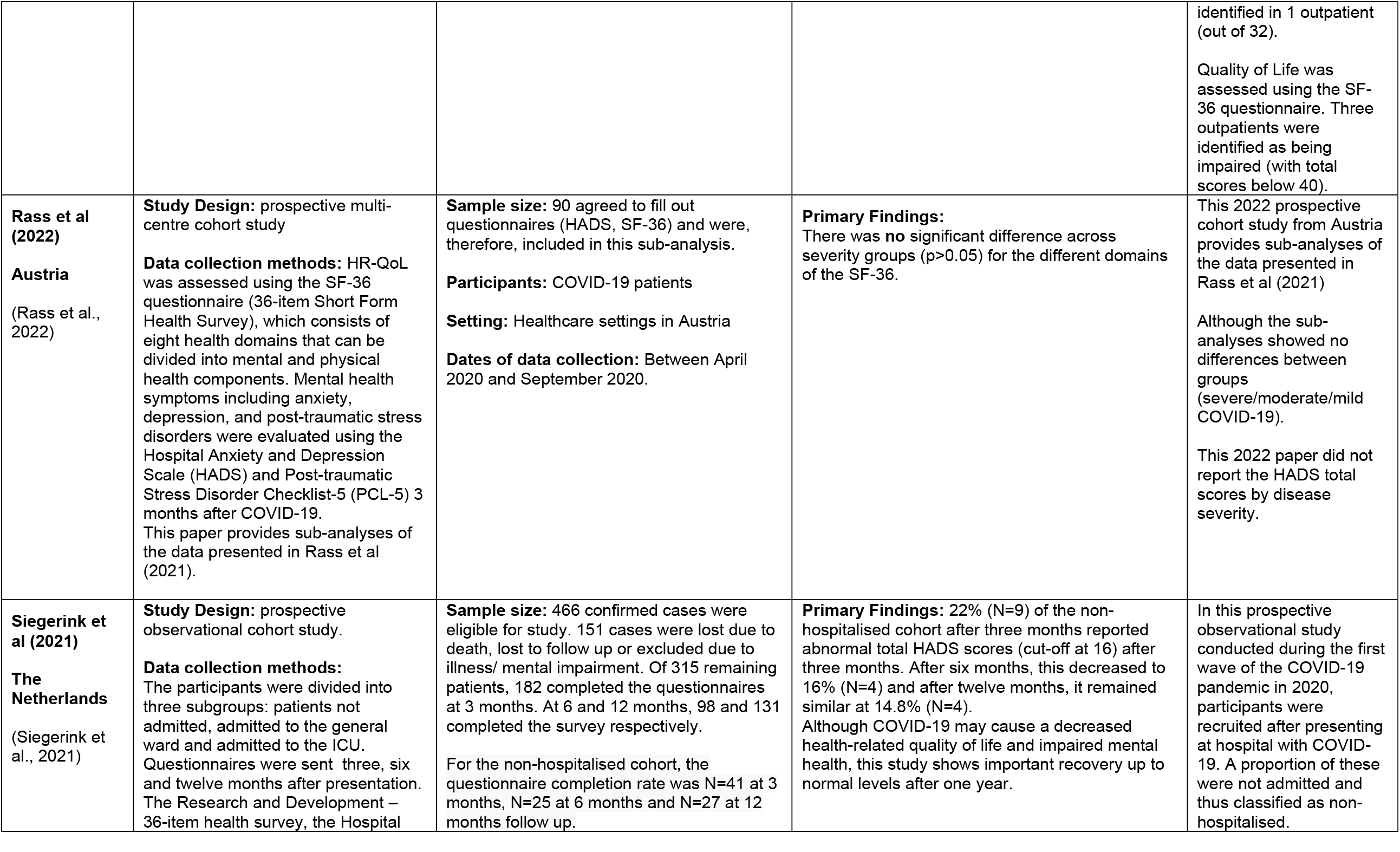
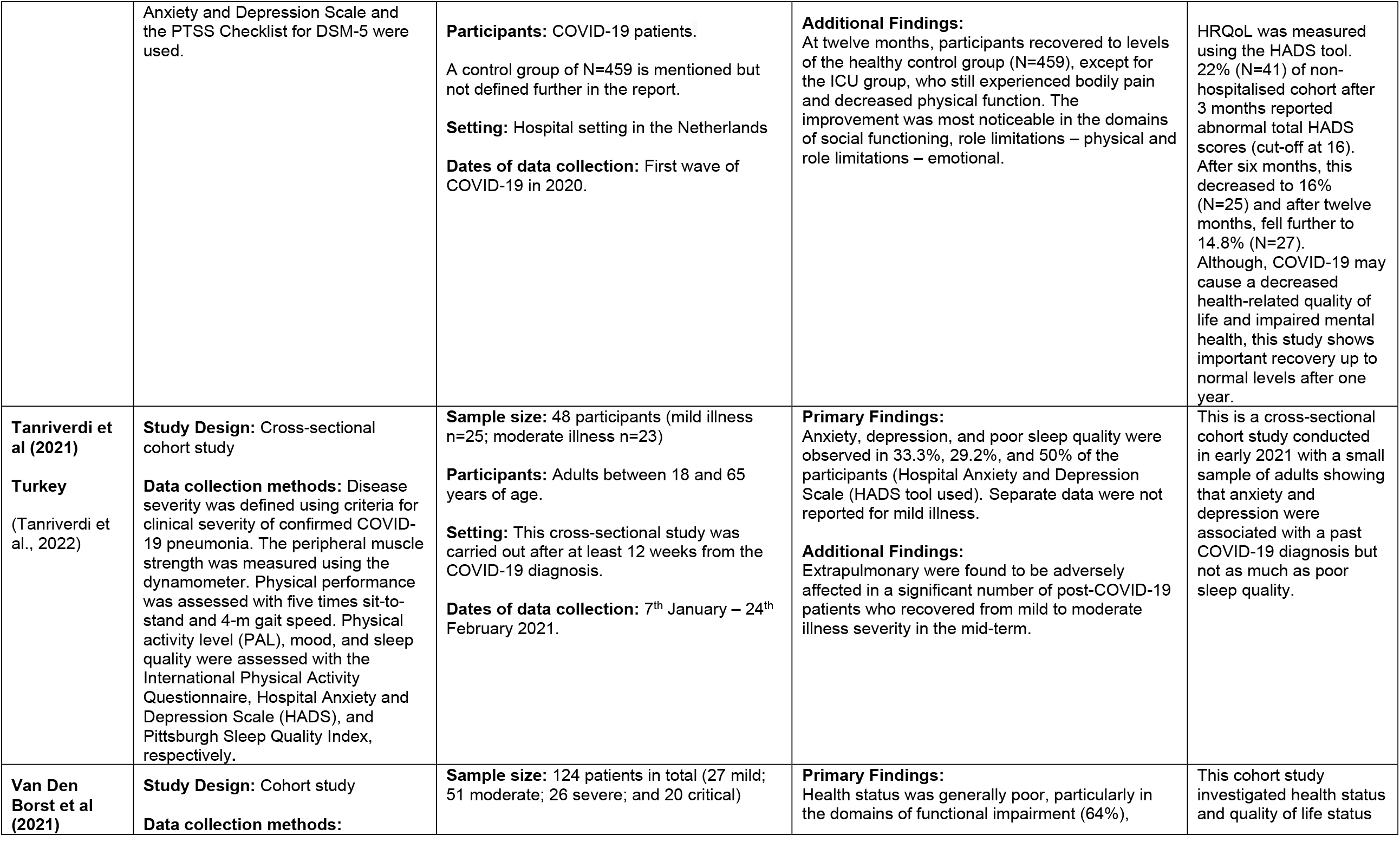
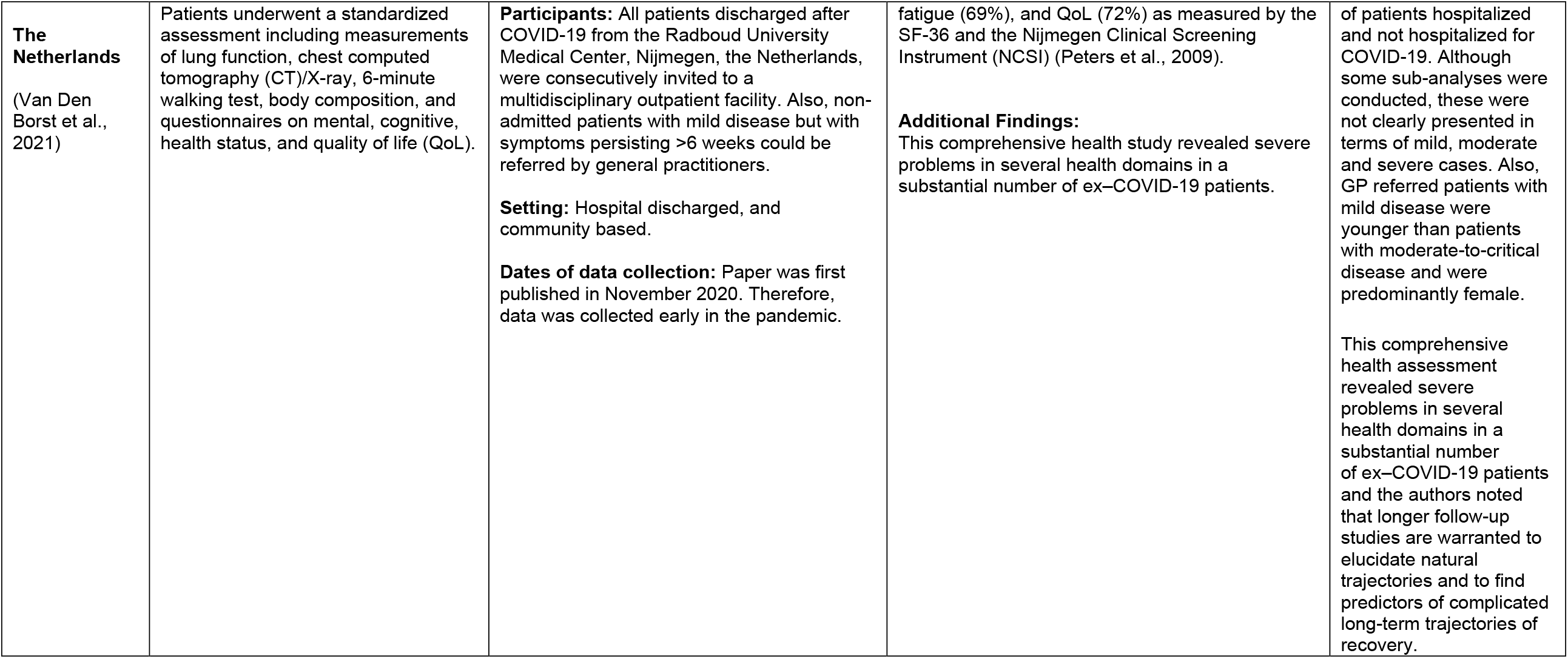
Primary studies – data extraction

| **Citation (Country)** | **Study Details** | **Participants & setting** | **Key findings** | **Observations/notes** |
| --- | --- | --- | --- | --- |
| **Akova and Gadikli 2022**<br>**(Turkey)**<br>(Akova & Gedikli, 2022) | **Study Design:** Cohort study<br>**Data collection methods:** Survey (demographic information and EQ-5D). The inclusion criteria for participation in the study were having previously been diagnosed with COVID-19 using the polymerase chain reaction (PCR) test and at least 12 weeks after negative test result. | **Sample size:** 151 adults. (141 outpatients and 10 inpatients)<br>**Participants:** 151 post-COVID-19 outpatients<br>**Setting:** Internal diseases outpatient clinic of Sivas Cumhuriyet University Hospital<br>**Dates of data collection:** March 2021 and April 2021 | **Primary Findings:**<br>Individuals with six or more post-COVID-19 symptoms had more problems with mobility ($p=0.012$), usual activities ($p=0.023$), pain/discomfort ($p=0.001$), and anxiety/depression ($p=0.002$) than individuals reporting zero or between 1 and 5 post-COVID-19 symptoms. The mean of the EuroQol Visual Analogue Scale (VAS) was also lower in individuals with six or more post-COVID-19 symptoms than in those who had no symptoms ($p=0.002$).<br>The mean duration of continued outpatient care after recovery from COVID-19 was $4.2 \pm 1.8$ months.<br>**Additional Findings:**<br>In female participants, post-COVID-19 symptoms increased by 2.7 times ($p=0.020$). The prevalence of certain COVID-19 symptoms was significantly higher in females (loss of smell, hair loss, and heart palpitation), those aged 40 years and older (intermittent fever), and obese individuals (heart palpitation and intermittent fever). | This cohort study from Turkey is important in terms of evaluating the ongoing symptoms after COVID-19, including the duration of symptoms, especially in terms of some sociodemographic characteristics of the participants, most of whom had completed their outpatient COVID-19 treatment. Considering that post-COVID-19 symptoms can be seen in patients with mild illness at a high rate and for a long period of time, patient follow-up should be given importance, especially for females and the elderly, as the more symptoms that persist, the lower the HRQoL. |
| **Attauabi et al 2022**<br>**(Denmark)**<br>(Attauabi et al., 2022) | **Study Design:** Cohort study<br>**Data collection methods:** This was a population-based study investigating the outcomes of COVID-19 among patients with Ulcerative Colitis and Crohn's disease (CD) in Denmark. Data was obtained through the Danish COVID-19 IBD Database, which monitors the disease course of laboratory-confirmed COVID-19 among patients with UC and CD. | **Sample size:** 222 patients with Inflammatory Bowel Diseases (IBD).<br>**Participants:** 137 patients with UC and 85 patients with CD. 125 patients experienced mild COVID-19 and 12 experienced adverse COVID-19.<br>**Setting:** IBD care setting<br>**Dates of data collection:** Unclear (pre-June 2022). | **Primary Findings:**<br>No difference was observed in terms of EQ-5D-5L among patients with UC and CD or patients with mild, adverse, or severe COVID-19.<br>**Additional Findings:**<br>CD patients with adverse COVID-19 experienced more fatigue than patients with mild COVID-19 (26 (IQR 25–35) vs. 41 (IQR 29–46), $p=0.03$). | According to this Danish cohort study, COVID-19 had no lasting impact on health-related quality of life among patients with Inflammatory Bowel Diseases (IBD) during the pandemic, providing further assurance for clinical guidelines during the pandemic for IBD care. |
|  | HRQoL was assessed at a median 5.1 months after COVID-19 infection. |  |  | The authors did not define the term 'adverse COVID-19'. |
| **Bileviciute-Ljungar et al 2022**<br>**Sweden**<br>(Bileviciute-Ljungar et al., 2022) | **Study Design:** Cohort study<br>**Data collection methods:** Survey methodology with a one-time, online questionnaire | **Sample size:** 106. 90% of patients reported that they were not-hospitalised during acute infection.<br>**Participants:** 82% female; 18% male<br>**Setting:** General Swedish population who had previous COVID-19 disease. Participants were recruited by Facebook sites and a stakeholder's organisation for post-COVID-19 syndrome in Sweden.<br>**Dates of data collection:** The data were collected from the end of April to the end of August 2021 (12 weeks after acute infection). | **Primary Findings:**<br>The number of persons with abnormal values of the EQ-5D and EQ-5D-VAS was n=99 and n=100, respectively. The mean for the EQ-5D was 0.51 (s.d. 0.2) and the range was 0.14-1.00.<br>On the EQ-5D-VAS the mean was 42.6 (s.d. = 19.5) and the range was 10-83.<br>**Additional Findings:**<br>The insomnia severity index (ISI), with a mean value of almost 13 was pathologically increased in 34% of participants. | The results of the present survey study show the complexity of the post-COVID-19 condition, indicating that pain is quite often reported by post-COVID-19 sufferers despite a mild initial infection.<br>The major limitation of this study is that those with pain were perhaps more inclined to complete a questionnaire on the topic. |
| **Han et al, 2022**<br>**USA**<br>(Han et al., 2022) | **Study Design:** Prospective cohort study<br>**Data collection methods:** Overall health status was measured with EQ-5D-5L and Patient Health Questionnaire-4 (PHQ-4). Data was obtained via electronic records and questionnaires across 3 time periods. Pre-illness baseline obtained from electronic records (1 month before positive infection) and peak acute COVID-19, and long-term follow-up (6– 11 months after the initial positive SARS-CoV-2 test) obtained via questionnaire. | **Sample size:** 436 outpatients<br>**Participants:** Outpatients with mild COVID-19. Of the 2092 outpatients with COVID-19, 436 (21%) responded to the survey.<br>**Setting:** Healthcare settings in the USA<br>**Dates of data collection:** April – July 2020 (6-11 months after acute COVID-19 disease) | **Primary Findings:**<br>Overall health status was measured by EQ-5D-VAS (0.63; 95% CI: 0.57–0.69), the EQ-5D-5L (0.65; 95%CI: 0.59–0.72), and psychological distress was measured by PHQ-4 (1.40; 95%CI, 1.28–1.54). | In this prospective cohort study from the USA in 2020 of participants with mild COVID-19, the burden of persistent symptoms was significantly associated with poorer long-term health status, poorer quality of life, and psychological distress. |
| **Labarca et al (2021)**<br>**Chile** | **Study Design:**<br>A prospective, cross-sectional, multi-centre study<br>**Data collection methods:** | **Sample size:** N=60 patients (n=18 mild, n=17 moderate, and n=25 severe) were include. | **Primary Findings**<br>For the (n=18) Mild COVID-19 patients, there was a mean SF-12 physical domain score at the 4-month follow-up of (50.3). Compared to mean scores of (37) and (41.2) for the moderate and severe patients, respectively. | In this multi-center study conducted in hospital and community settings in Chile from April to June 2020, it was found that patients diagnosed with |
| (Labarca et al., 2021) | Patients with different illness severities were consecutively included (mild; moderate: hospitalised without invasive mechanical ventilation (IMV); severe: hospitalised with IMV). Clinical variables, health-related quality of life (HRQoL) assessed via the Short Form 12 Health Survey (SF-12), the latter used to evaluate HRQoL and included both physical and mental domains.<br>Baseline data was extracted detailing a patient's previous medical records and COVID-19 illness during the acute phase.<br>After four months, a follow-up was conducted. Participants filled out SF-12 and HADS questionnaires. | **Participants:** Patients aged $\geq 18$ years with previous positive PCR for SARS-CoV-2 infection from April to July 2020.<br>**Setting:** Hospital and community settings in Chile.<br>**Dates of data collection:** April to July 2020. | HRQoL was measured using SF-12. Personal change in HRQoL was then assessed using a visual analogue scale with a range of 0% (worst HRQoL) and 100% (best HRQoL) at baseline (before SARS-CoV-2 infection) and during the follow-up. A change of $\geq 10\%$ was indicative of a change in HRQoL.<br>50% of the (n=18) mild COVID-19 patients reported an individual change in HRQoL of $\geq 10\%$.<br>The mean HADS-A (Anxiety) score for the mild group was 5.87.<br>The mean HADS-D (Depression) score for the mild group was 5.5.<br>In conclusion, patients diagnosed with COVID-19 presented a high prevalence of symptoms and impaired quality of life, regardless of infection severity.<br>**Additional findings**<br>After four months of follow-up, the prevalence of severe fatigue is frequent among patients with different severities of COVID-19 illness. | COVID-19 presented a high prevalence of symptoms and impaired quality of life (as measured by the HADS) regardless of infection severity.<br>Mild COVID-19 was defined in this study as mild symptoms (e.g., fever, cough, and change in taste or smell, without dyspnea). Mild cases were determined according to $\text{PaO}_2/\text{FiO}_2 > 250$ when measured and when the patient reported mild symptoms (eg, fever, cough, and change in taste or smell) without dyspnea according to the initial telephonic evaluation and during follow-up clinical evaluation.<br>The Mild COVID-19 group received clinical outpatient monitoring and supportive care, and results for this group only are included in this table. |
| **Mayer et al (2021)**<br>**USA**<br>(Mayer et al., 2021) | **Study Design:** Case report.<br>**Type of intervention [exposure]:** Bi-weekly sessions for eight weeks (16 sessions) of physical therapist treatment starting 64 days after their initial positive test. This included aerobic training, strengthening exercises, diaphragmatic breathing | **Sample size:** 1<br>**Participants:** A 37-year-old woman who tested positive for SARS-CoV-2 and developed mild COVID-19 disease but did not require supplemental oxygen or hospitalisation<br>**Setting:** Pulmonary rehabilitation clinic. | **Primary Findings:**<br>The EQ-5D VAS measure was used to investigate health-related quality of life at two time points. At baseline (day 62 after initial positive diagnosis of COVID-19), the EQ-5D VAS score was 50. At follow-up (after eight weeks of physical therapy treatment (approx. 120 days after initial positive COVID-19 diagnosis)), the EQ-5D VAS score decreased to 40). | This case report from the USA conducted prior to publication in 2021, found that quality of life scores measured by EQ-5D VAS and PTSD scores did not improve following eight weeks of physiotherapy. This was |
|  | techniques, mindfulness training, and education.<br>**Data collection methods:** Physical therapist evaluation and self-reported symptoms. | **Dates of data collection:** Prior to publication of paper in 2021. | **Additional Findings:**<br>Following physiotherapy, the patient's muscle strength, physical function, and exercise capacity improved. 6-Minute walk distance increased by 199 m, equating to 80% of their age-predicted distance.<br>At evaluation after physical therapy, the patient was still experiencing migraines, dyspnoea, fatigue, and cognitive dysfunction. | despite improvements in physical functioning. |
| **Matviyets and Matiukha (2022)**<br>Ukraine<br>(Matviyets & Matiukha, 2022) | **Study Design:** Cohort study<br>**Data collection methods:** Electronic outpatient cards. PHQ 9 screening for post-traumatic stress disorder (PTSD). | **Sample size:** 70 (n=23 with mild COVID-19)<br>**Participants:** COVID-19 Outpatients aged from 31 to 80 years<br>**Setting:** Health care setting in Ukraine.<br>**Dates of data collection:** Collection date not defined. | **Primary Findings:**<br>52% of the sample who had mild COVID-19 (n=23) reported anxiety as measured by the PHQ 9 tool.<br>**Additional Findings:**<br>Of the mild COVID-19 sample n=23, 26% had PTSD, 26% reported disorders of cognitive functions, 83% had sleep disorders and 52% had Tachycardia. | This cohort study from Ukraine assessed the mental health of both inpatients and outpatients who had confirmed diagnosis of SARS CoV-2. They found that 52% of the sample with mild COVID-19 reported anxiety as measured by the PHQ 9 tool. |
| **Ordinola Navarro et al (2021)**<br>Mexico<br>(Ordinola Navarro et al., 2021) | **Study Design:** Cohort study<br>**Data collection methods:** Survey and observational methods. Baseline data was obtained via patient self-reporting of general health condition before COVID-19 infection. | **Sample size:** (N=115). N=43 classified as Mild, n=38 classified as moderate and n=34 classified as severe and critically ill.<br>**Participants:** Patients evaluated for COVID-19 in The Hospital de Especialidades "Dr. Antonio Fraga Mouret," Centro Medico Nacional La Raza, Mexico.<br>**Setting:** Healthcare settings in Mexico.<br>**Dates of data collection:** between April and June 2020 (at least 30 days post COVID-19) | **Primary Findings:**<br>The median score of the EQ-5D-5L visual analogue scale pre-COVID-19 was 95 (IQR; 90–100), and after was 85 (IQR; 75–90), $P < 0.001$. (EQ-5D scores were not presented according to severity of initial COVID-19 infection).<br>Of the n=43 mild patients, 22 reported a severe decrease in QoL. A severe decrease in QoL was defined as a decrease of $\geq 10$ points in the EQ-5D-5L. 21 reported no decrease in QoL.<br>Of the n=38 moderate patients, 25 reported a severe decrease in QOL. 13 reported no decrease in QOL. A severe decrease in QOL is defined as a decrease of at least 10 points in the EQ-5d-5L visual analogue scale. | A cohort study based in healthcare settings in Mexico investigated the HRQoL of those with mild/Severe/critically ill with COVID-19 in 2020. It was found that even those with mild symptoms showed a decrease in quality of life as measured by the EQ-5D VAS. Scores on the EQ-5D showed that mobility, anxiety/depression and usual activities were affected more than self-care. |
|  |  |  | In this follow-up study of recovered COVID-19 patients, the authors found a high proportion of patients with a decrease in QoL, and this finding seemed not to have a relation with the disease severity and the persistence of symptoms. Moreover, alterations in health parameters such as usual activities and anxiety/depression were present.<br>**Additional Findings:**<br>Across the whole cohort (all severity classifications) the EQ-5D-5L domains with the greatest difference pre and post-COVID-19 were Pain/discomfort, Mobility and Anxiety/depression. Usual activities also saw a difference between pre and post-COVID-19, and the Self-care domain saw little change. | This study may be limited in the fact that pre-COVID HRQoL was determined through patient self-reporting of their perceived health condition before COVID-19 infection. Furthermore, the study provides limited long-term follow-up at 30 days. |
| **Rass et al (2021)**<br>**Austria**<br>(Rass et al., 2021) | **Study Design:** prospective, multi-centre, observational cohort study.<br>**Data collection methods:**<br>HRQoL was assessed using the SF-36 questionnaire (36-item Short Form Health Survey), which consists of eight health domains that can be divided into mental and physical health components. Mental health symptoms, including anxiety, depression, and post-traumatic stress disorders were evaluated using the Hospital Anxiety and Depression Scale (HADS) and Post-traumatic Stress Disorder Checklist-5 (PCL-5) 3 months after COVID-19. COVID-19 patients presenting at hospital were prospectively enrolled during the acute phase of the disease.<br>Baseline was defined as the day of diagnosis by a positive SARSCoV-2 test result. | **Sample size:** 135 patients; 32 (24%) patients were treated as outpatients (non-hospitalised).<br>A total of 135/145 (93%) patients agreed to participate in the neurological follow-up three months after disease onset and were evaluated using a structured neurological assessment.<br>**Participants:** COVID-19 patients.<br>**Setting:** Healthcare settings in Austria<br>**Dates of data collection:** Between April 2020 and September 2020. | **Primary Findings:**<br>Depression was assessed utilising the HADS-D. HADS-D scores of >7 was identified in 1 outpatient (out of 32). A HADS-D score of >10 was identified in 1 outpatient (out of 32).<br>Anxiety assessed using the HADS-A. A HADS-A score of >7 was identified in 5 outpatients (out of 32). A HADS-A score of >10 was identified in 1 outpatient (out of 32).<br>Quality of Life was assessed using the SF-36 questionnaire. Three outpatients were identified as being impaired (with total scores below 40).<br>Eight outpatients suffered with persistent fatigue and nine reported sleep disturbances.<br>**Additional Findings:**<br>PTSD identified (Categorised as a score of >32 on the PCL) was reported in 1 outpatient (out of 32). | An Austrian study conducted in 2020 included participants recruited after presenting at hospital. A proportion of these were treated as outpatients and thus classified as non-hospitalised.<br>Depression was assessed utilising the HADS-D. HADS-D scores of >7 was identified in 1 outpatient (out of 32). A HADS-D score of >10 was identified in 1 outpatient (out of 32).<br>Anxiety was assessed using the HADS-A. A HADS-A score of >7 was identified in 5 outpatients (out of 32). A HADS-A score of >10 was<br>identified in 1 outpatient (out of 32).<br>Quality of Life was assessed using the SF-36 questionnaire. Three outpatients were identified as being impaired (with total scores below 40). |
| **Rass et al (2022)**<br>**Austria**<br>(Rass et al., 2022) | **Study Design:** prospective multi-centre cohort study<br>**Data collection methods:** HR-QoL was assessed using the SF-36 questionnaire (36-item Short Form Health Survey), which consists of eight health domains that can be divided into mental and physical health components. Mental health symptoms including anxiety, depression, and post-traumatic stress disorders were evaluated using the Hospital Anxiety and Depression Scale (HADS) and Post-traumatic Stress Disorder Checklist-5 (PCL-5) 3 months after COVID-19. This paper provides sub-analyses of the data presented in Rass et al (2021). | **Sample size:** 90 agreed to fill out questionnaires (HADS, SF-36) and were, therefore, included in this sub-analysis.<br>**Participants:** COVID-19 patients<br>**Setting:** Healthcare settings in Austria<br>**Dates of data collection:** Between April 2020 and September 2020. | **Primary Findings:**<br>There was **no** significant difference across severity groups ($p>0.05$) for the different domains of the SF-36. | This 2022 prospective cohort study from Austria provides sub-analyses of the data presented in Rass et al (2021)<br>Although the sub-analyses showed no differences between groups (severe/moderate/mild COVID-19).<br>This 2022 paper did not report the HADS total scores by disease severity. |
| **Siegerink et al (2021)**<br>**The Netherlands**<br>(Siegerink et al., 2021) | **Study Design:** prospective observational cohort study.<br>**Data collection methods:** The participants were divided into three subgroups: patients not admitted, admitted to the general ward and admitted to the ICU. Questionnaires were sent three, six and twelve months after presentation. The Research and Development – 36-item health survey, the Hospital | **Sample size:** 466 confirmed cases were eligible for study. 151 cases were lost due to death, lost to follow up or excluded due to illness/ mental impairment. Of 315 remaining patients, 182 completed the questionnaires at 3 months. At 6 and 12 months, 98 and 131 completed the survey respectively.<br>For the non-hospitalised cohort, the questionnaire completion rate was N=41 at 3 months, N=25 at 6 months and N=27 at 12 months follow up. | **Primary Findings:** 22% (N=9) of the non-hospitalised cohort after three months reported abnormal total HADS scores (cut-off at 16) after three months. After six months, this decreased to 16% (N=4) and after twelve months, it remained similar at 14.8% (N=4). Although COVID-19 may cause a decreased health-related quality of life and impaired mental health, this study shows important recovery up to normal levels after one year. | In this prospective observational study conducted during the first wave of the COVID-19 pandemic in 2020, participants were recruited after presenting at hospital with COVID-19. A proportion of these were not admitted and thus classified as non-hospitalised. |
|  | Anxiety and Depression Scale and the PTSS Checklist for DSM-5 were used. | **Participants:** COVID-19 patients.<br>A control group of N=459 is mentioned but not defined further in the report.<br>**Setting:** Hospital setting in the Netherlands<br>**Dates of data collection:** First wave of COVID-19 in 2020. | **Additional Findings:**<br>At twelve months, participants recovered to levels of the healthy control group (N=459), except for the ICU group, who still experienced bodily pain and decreased physical function. The improvement was most noticeable in the domains of social functioning, role limitations – physical and role limitations – emotional. | HRQoL was measured using the HADS tool. 22% (N=41) of non-hospitalised cohort after 3 months reported abnormal total HADS scores (cut-off at 16). After six months, this decreased to 16% (N=25) and after twelve months, fell further to 14.8% (N=27). Although, COVID-19 may cause a decreased health-related quality of life and impaired mental health, this study shows important recovery up to normal levels after one year. |
| **Tanriverdi et al (2021)**<br>**Turkey**<br>(Tanriverdi et al., 2022) | **Study Design:** Cross-sectional cohort study<br>**Data collection methods:** Disease severity was defined using criteria for clinical severity of confirmed COVID-19 pneumonia. The peripheral muscle strength was measured using the dynamometer. Physical performance was assessed with five times sit-to-stand and 4-m gait speed. Physical activity level (PAL), mood, and sleep quality were assessed with the International Physical Activity Questionnaire, Hospital Anxiety and Depression Scale (HADS), and Pittsburgh Sleep Quality Index, respectively. | **Sample size:** 48 participants (mild illness n=25; moderate illness n=23)<br>**Participants:** Adults between 18 and 65 years of age.<br>**Setting:** This cross-sectional study was carried out after at least 12 weeks from the COVID-19 diagnosis.<br>**Dates of data collection:** 7<sup>th</sup> January – 24<sup>th</sup> February 2021. | **Primary Findings:**<br>Anxiety, depression, and poor sleep quality were observed in 33.3%, 29.2%, and 50% of the participants (Hospital Anxiety and Depression Scale (HADS tool used). Separate data were not reported for mild illness.<br>**Additional Findings:**<br>Extrapulmonary were found to be adversely affected in a significant number of post-COVID-19 patients who recovered from mild to moderate illness severity in the mid-term. | This is a cross-sectional cohort study conducted in early 2021 with a small sample of adults showing that anxiety and depression were associated with a past COVID-19 diagnosis but not as much as poor sleep quality. |
| **Van Den Borst et al (2021)** | **Study Design:** Cohort study<br>**Data collection methods:** | **Sample size:** 124 patients in total (27 mild; 51 moderate; 26 severe; and 20 critical) | **Primary Findings:**<br>Health status was generally poor, particularly in the domains of functional impairment (64%), | This cohort study investigated health status and quality of life status |
| **The Netherlands**<br>(Van Den Borst et al., 2021) | Patients underwent a standardized assessment including measurements of lung function, chest computed tomography (CT)/X-ray, 6-minute walking test, body composition, and questionnaires on mental, cognitive, health status, and quality of life (QoL). | **Participants:** All patients discharged after COVID-19 from the Radboud University Medical Center, Nijmegen, the Netherlands, were consecutively invited to a multidisciplinary outpatient facility. Also, non-admitted patients with mild disease but with symptoms persisting >6 weeks could be referred by general practitioners.<br>**Setting:** Hospital discharged, and community based.<br>**Dates of data collection:** Paper was first published in November 2020. Therefore, data was collected early in the pandemic. | fatigue (69%), and QoL (72%) as measured by the SF-36 and the Nijmegen Clinical Screening Instrument (NCSI) (Peters et al., 2009).<br>**Additional Findings:**<br>This comprehensive health study revealed severe problems in several health domains in a substantial number of ex-COVID-19 patients. | of patients hospitalized and not hospitalized for COVID-19. Although some sub-analyses were conducted, these were not clearly presented in terms of mild, moderate and severe cases. Also, GP referred patients with mild disease were younger than patients with moderate-to-critical disease and were predominantly female.<br>This comprehensive health assessment revealed severe problems in several health domains in a substantial number of ex-COVID-19 patients and the authors noted that longer follow-up studies are warranted to elucidate natural trajectories and to find predictors of complicated long-term trajectories of recovery. |

### 6.3 Information available on request

All the papers referenced in this Rapid Review are available from the relevant journals.

## 7. ADDITIONAL INFORMATION

## 7.2 Acknowledgements

The authors would like to thank expert stakeholders from the Department of Health and Social Care (DHSC), including, Mina Mahmoudzadeh (Research analyst in the COVID-19 Health Protection Analysis Team at DHSC), Gbemi Babalola (Economic Adviser at Department of Health and Social Care), Claire Bradshaw (Analyst at Department for Health and Social Care) and Marianne Scholes (Senior Analyst at Department for Health and Social Care). From the Welsh Government’s Technical Advisory Cell, Dr Brendan Collins (Senior Health Economist). We thank Professor Adrian Edwards, Dr Ruth Lewis and Dr Alison Cooper (Wales COVID-19 Evidence Centre) for providing support to the review team. Dr Catherine Lawrence is also thanked for lending her professional knowledge in ensuring the factual correctness of the details and for proofreading the report.

## 7.3 Author contributions

Contribution to writing and critical editing of the report; LHS, AH, KP, JD, AM, CW, RTE, DH, DF.

## 7.4 Disclaimer

The views expressed in this publication are those of the authors, not necessarily Health and Care Research Wales. The WCEC and authors of this work declare that they have no conflict of interest.

## 8. ABOUT THE WALES COVID-19 EVIDENCE CENTRE (WCEC)

The WCEC integrates with worldwide efforts to synthesise and mobilise knowledge from research.

We operate with a core team as part of <u>Health and Care Research Wales</u>, are hosted in the <u>Wales Centre for Primary and Emergency Care Research (PRIME),</u> and are led by <u>Professor Adrian Edwards of Cardiff University.</u>

The core team of the centre works closely with collaborating partners in<u>Health Technology Wales</u>, <u>Wales Centre for Evidence-Based Care</u>, <u>Specialist Unit for Review Evidence centre</u>, <u>SAIL Databank</u>, <u>Bangor Institute for Health & Medical Research/ Health and Care Economics Cymru,</u> and the <u>Public Health Wales Observatory.</u>

Together we aim to provide around 50 reviews per year, answering the priority questions for policy and practice in Wales as we meet the demands of the pandemic and its impacts.

### Director

Professor Adrian Edwards

### Website

https://healthandcareresearchwales.org/about-research-community/wales-covid-19-evidence-centre

## APPENDICES

**Appendix 1:** Description of the Health-Related Quality of Life (HRQoL) measures.

| # | Name of HRQoL measure | Description of HRQoL measure | Application | Link to relevant website |
| --- | --- | --- | --- | --- |
| 1. | EQ-5D (EuroQol Group, 1990) | The EQ-5D is an instrument which evaluates generic quality of life. It was developed in Europe and is widely used. The EQ-5D descriptive system is a preference-based HRQoL measure with one question for each of the following five dimensions: mobility, self-care, usual activities, pain/discomfort, and anxiety/depression.<br>Health states can be given a summary index score based on societal preferences. Health state index scores generally range from less than 0 (where 0 is a health state equivalent to death; negative values are valued as worse than death) to 1 (perfect health), with higher scores indicating higher health utility, though health state preferences can differ between countries (EuroQol Research Foundation, 2018, 2019). | The EQ-5D measure is widely used in research and in surgical settings to measure HRQoL. | https://euroqol.org/support/how-to-obtain-eq-5d/ |
| 2. | Hospital Anxiety and Depression Scale (HADS) (Zigmond & Snaith, 1983) | The HADS self-assessment scale was developed in the early 1980s and has been found to be a reliable instrument for detecting states of depression and anxiety in the setting of a hospital medical outpatient clinic. The anxiety and depressive subscales are also valid measures of severity of the emotional disorder. The HADS is a 14-question instrument that measures anxiety and depression. Each question is scored between 0 (no impairment) and 3 (severe impairment), with a maximum score of 21 for anxiety or depression. The HADS itself is not explicitly preference-based, but it can be mapped to the EQ-5D to generate a cost per quality adjusted life year (QALY). | The HADS measure is widely used in research and in surgical settings to measure HRQoL. | https://www.svri.org/sites/default/files/attachments/2016-01-13/HADS.pdf |
| 3. | Patient Health Questionnaire-9 (Kroenke & Spitzer, 2002) | The PHQ-9 is a depression scale, which is half the length of many other depression measures, has comparable sensitivity and specificity, and consists of the actual nine criteria on which the diagnosis of DSM-IV depressive disorders is based. The latter feature distinguishes the PHQ-9 from other two-step depression measures for which, when scores are high, additional questions must be asked to establish DSM-IV depressive diagnoses. The PHQ-9 is thus a dual-purpose instrument that, with the | The PHQ-9 has been validated for use in primary care. It is not a screening tool for depression, but it is used to monitor the severity of depression and response to treatment. | https://patient.info/doctor/patient-health-questionnaire-phq-9 |
|  |  | same nine items, can establish provisional depressive disorder diagnoses as well as grade depressive symptom severity.<br>The PHQ-9 itself is not explicitly a preference-based tool, but it can be mapped to the SF-6D (Brazier et al., 2002) to generate a cost per quality adjusted life year (QALY). | The PHQ-4 is a short form of the PHQ-9. |  |
| 4. | SF-36 (and shorter version, SF-12) (Ware, 2000) | The SF-36 is a multipurpose, short-form health survey with 36 questions. It yields an eight-scale profile of scores as well as physical and mental health summary measures. It is a generic measure, as opposed to one that targets a specific age, disease, or treatment group. Accordingly, the SF-36 has been useful in comparing general and specific populations, comparing the relative burden of diseases, differentiating the health benefits produced by a wide range of different treatments, and screening individual patients. The SF-12 is a shorter version of the short-form 36.<br>The SF-36 and the SF-12 can be used to generate a preference-based index via the SF-6D (Brazier et al., 2002). | The SF-36 and SF-12 are widely used in research and clinical practice. | https://clinmedjournals.org/articles/jmdt/jmdt-2-023-figure-1.pdf |

**Appendix 2:**
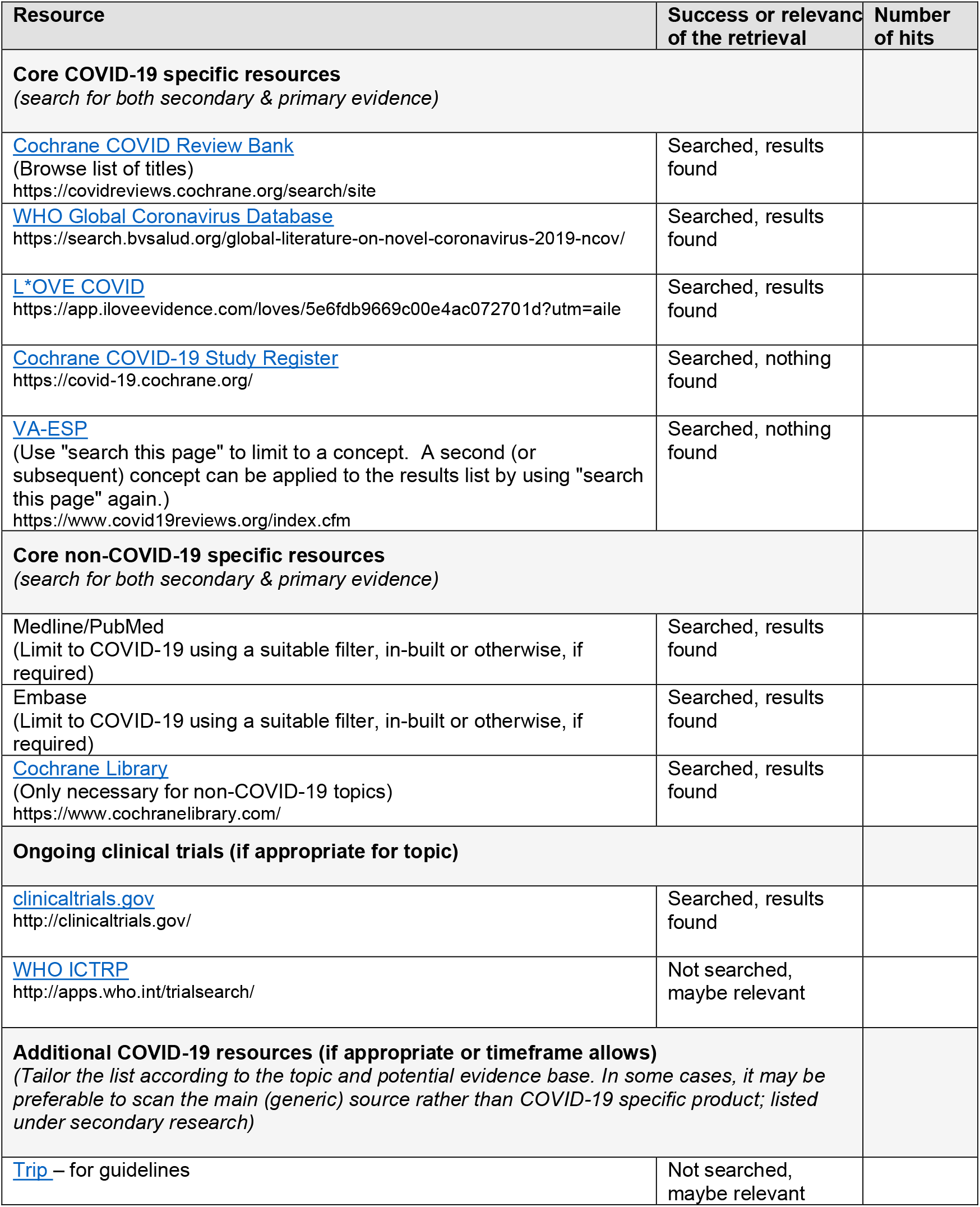
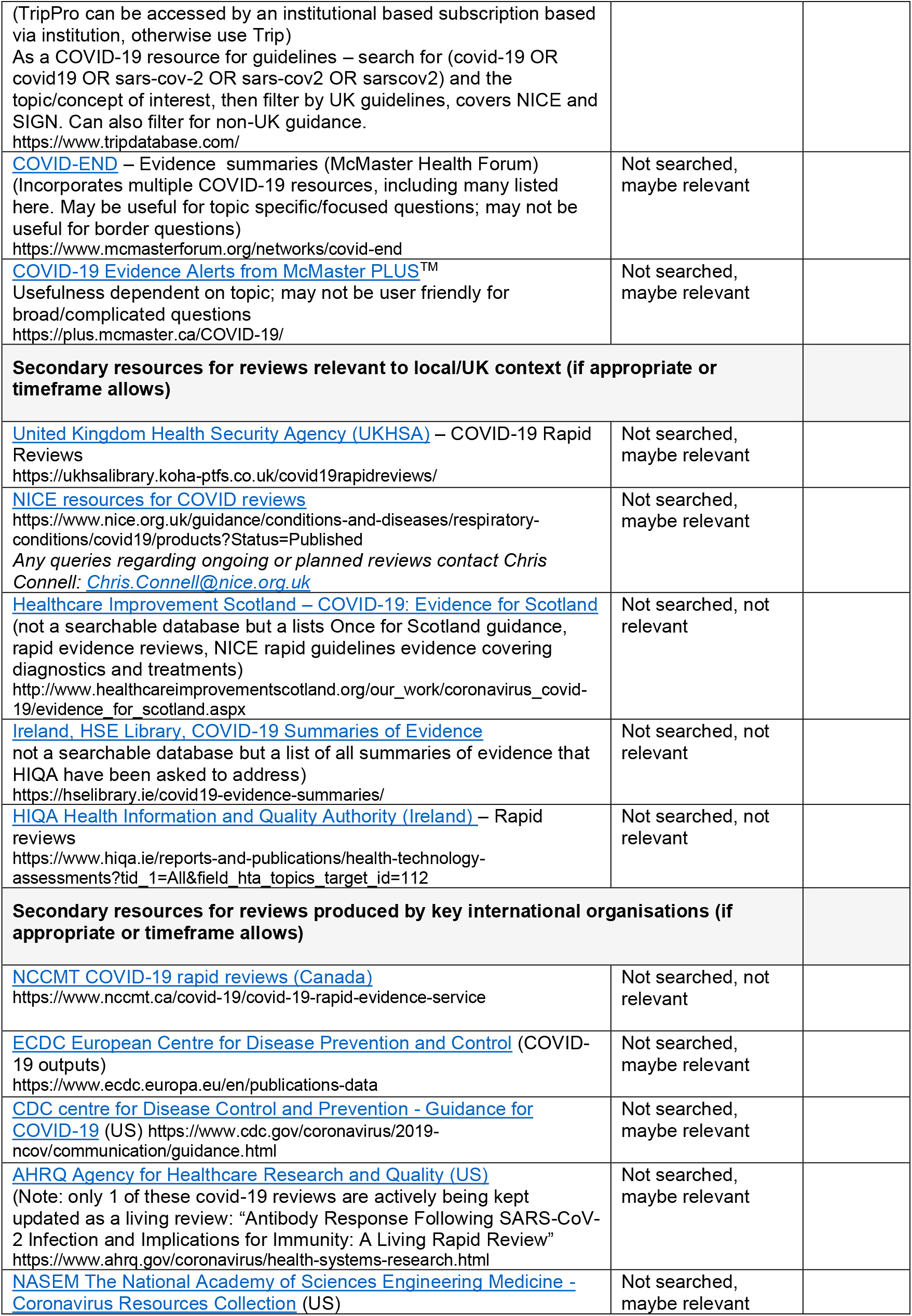
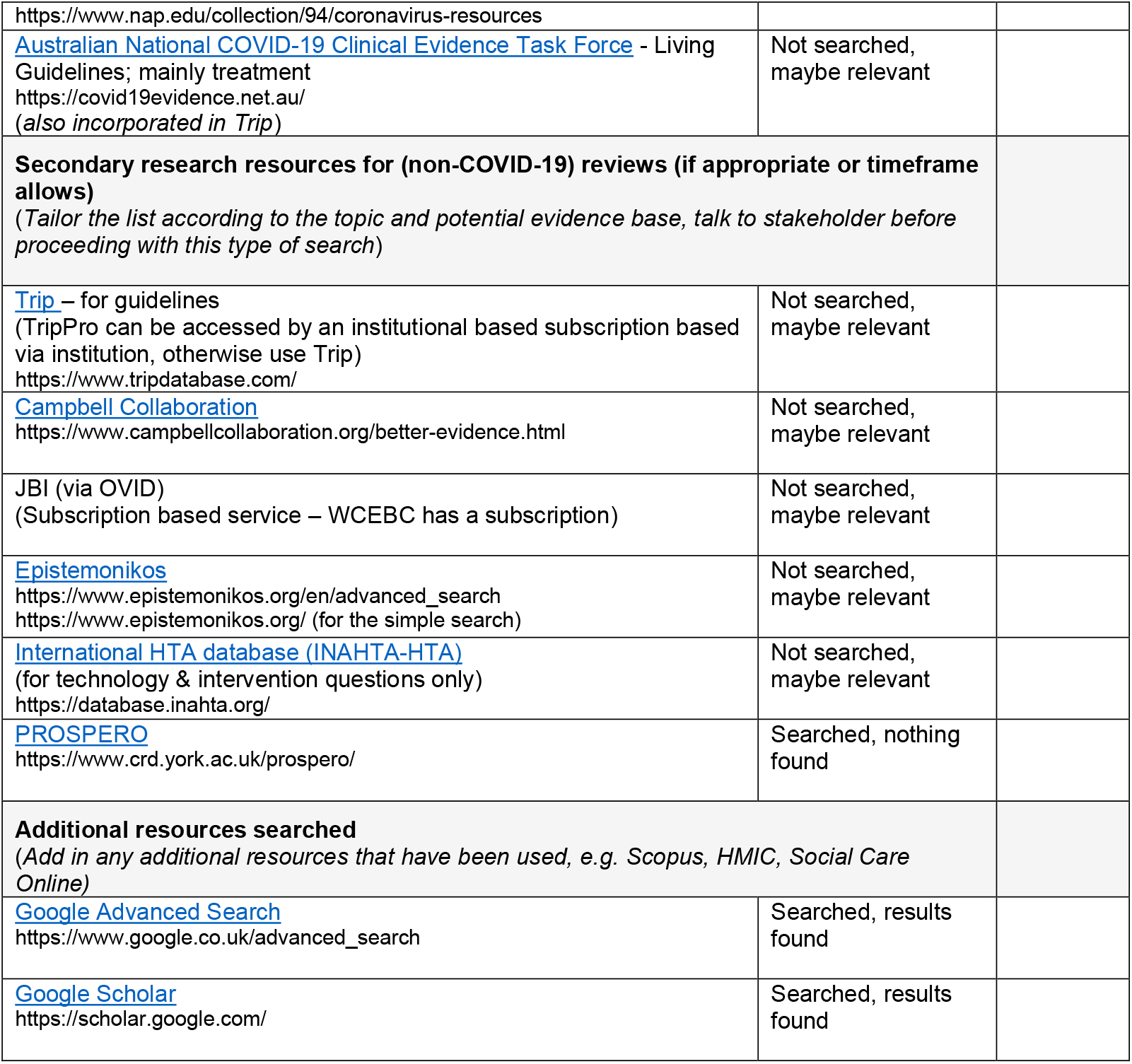
Resources searched during Rapid Review Searching. A single list of resources has been developed for guiding and documenting the sources searched as part of a Rapid Review. All ‘core’ resources should be searched, but other resources may be considered if appropriate to the topic, or time allows. For those resources used, record the search strategies used below the table.

**Appendix 3:** Map of Mild COVID-19 and HRQoL by type of evidence. **Map of the evidence COVID-19 and Health Related Quality of Life (HRQoL)**

| Type of evidence | Mild COVID-19 disease and HRQoL studies by year published |  |  | Total |
| --- | --- | --- | --- | --- |
|  | 2020 | 2021 | 2022 |  |
| Systematic Review (SR) | 0 | Poudel et al (2021) | Bourmistrova et al (2022) | 2 |
| Cohort study | 0 | Labarca et al (2021) | Akova and Gadikli et al (2022) |  |
|  |  | Ordinola Navarro et al (2021) | Attauabi et al (2022) |  |
|  |  | Rass et al (2021) | Bileviciute-Ljungar et al (2022) |  |
|  |  | Siegerink et al (2021) | Han et al (2022) |  |
|  |  | Van Den Borst et al (2021) | Matviyets & Matiukha (2022) |  |
|  |  |  | Rass et al (2022) |  |
|  |  |  | Tanriverdi et al (2022) | 12 |
| Case report | 0 | Mayer et al (2021) | 0 | 1 |
| **Total** | **0** | **7** | **8** | **15** |

**Appendix 4: Quality appraisal tables**

Members of the review team chose the most appropriate JBI critical appraisal tool to quality appraise the included studies. (See Tables A.4.1-A.4.3 below).

**Table A.4.1.** JBI cohort checklist.

| Citation | Q1. Were the two groups similar and recruited from the same population? | Q2. Were the exposures measured similarly to assign people to both exposed and unexposed groups? | Q3. Was the exposure measured in a valid and reliable way? | Q4. Were confounding factors identified? | Q5. Were strategies to deal with confounding factors stated? | Q6. Were the groups/ participants free of the outcome at the start of the study (or at the moment of exposure)? | Q7. Were the outcomes measured in a valid and reliable way? | Q8. Was the follow up time reported and sufficient to be long enough for outcomes to occur? | Q9. Was follow up complete, and if not, were the reasons to loss to follow up described and explored? | Q10. Were strategies to address incomplete follow up utilized? | Q11. Was appropriate statistical analysis used? |
| --- | --- | --- | --- | --- | --- | --- | --- | --- | --- | --- | --- |
| Akova and Gadikli (2022)<br>* | Yes | Yes | Yes | No | No | No | Yes | N/A | N/A | N/A | Yes |
| Attauabi et al (2022) | Yes | Yes | Yes | No | No | Yes | Yes | No | No | No | Yes |
| Bileviciute-Ljungar (2022) | Yes | Yes | Yes | Yes | Yes | Yes | Yes | N/A | N/A | N/A | Yes |
| Han et al, 2022 | Yes | Yes | Yes | Yes | Yes | Yes | Yes | Yes | Yes | N/A | Yes |
| Labarca et al., 2021 | Yes | Yes | Yes | Yes | Yes | No | Yes | Yes | No | No | Yes |
| Matviyets et al, 2022 | Yes | Yes | Unclear | Yes | Yes | Yes | Yes | No | Unclear | N/A | Yes |
| Ordinola Navarro et al., 2021 | Yes | Yes | Yes | Yes | Yes | No | Yes | Yes | Yes | No | Yes |
| Rass et al., 2021 | Yes | Yes | Yes | Yes | Yes | No | Yes | Yes | Yes | No | Yes |
| Rass et al., 2022 | Yes | Yes | Yes | Yes | Yes | No | Yes | Yes | Yes | No | Yes |
| Siegerink et al., 2021 | Yes | Yes | Yes | Yes | No | No | Yes | Yes | Yes | No | Yes |
| Tanriverdi et al (2021) | Yes | Yes | Yes | Yes | Yes | Yes | Yes | No | No | No | Yes |
| Van Den Borst et al (2021) | Yes | Yes | Yes | No | No | Yes | Yes | Yes | Yes | Yes | Yes |

**Table A.4.2.** JBI Systematic Reviews and Research Syntheses Checklist.

| Citation |  | Q1. Is the review question clearly and explicitly stated? | Q2. Were the inclusion criteria appropriate for the review question? | Q3. Was the search strategy appropriate? | Q4. Were the sources and resources used to search for studies adequate? | Q5. Were the criteria for appraising studies appropriate? | Q6. Was critical appraisal conducted by two or more reviewers independently? | Q7. Were there methods to minimize errors in data extraction? | Q8. Were the methods used to combine studies appropriate? | Q9. Was the likelihood of publication bias assessed? | Q10. Were recommendations for policy and/or practice supported by the reported data? | Q11. Were the specific directives for new research appropriate? |
| --- | --- | --- | --- | --- | --- | --- | --- | --- | --- | --- | --- | --- |
| Bourmistrova et al (2022) |  | Yes | Yes | Yes | Yes | Yes | Yes | Yes | Yes | Yes | Yes | Unclear |
| Poudel et al (2021) |  | Unclear | Yes | Yes | Yes | Yes | Yes | Yes | Yes | Yes | Yes | Yes |

**Table A.4.3.** JBI Critical Appraisal Checklist for Case Reports.

| Citation | Q1. Were patient's demographic characteristics clearly described? | Q2. Was the patient's history clearly described and presented as a timeline? | Q3. Was the current clinical condition of the patient on presentation clearly described? | Q4. Were diagnostic tests or assessment methods and the results clearly described? | Q5. Was the intervention(s) or treatment procedure(s) clearly described? | Q6. Was the post-intervention clinical condition clearly described? | Q7. Were adverse events (harms) or unanticipated events identified and described? | Q8. Does the case report provide takeaway lessons? |
| --- | --- | --- | --- | --- | --- | --- | --- | --- |
| Mayer et al, 2021 | Yes | Yes | Yes | Yes | Yes | Yes | No | Yes |

